# A Randomized Controlled Trial to Increase Cancer Screening and Reduce Depression among Low-Income Women

**DOI:** 10.1101/2020.11.05.20225383

**Authors:** Jonathan N. Tobin, Elisa S. Weiss, Andrea Cassells, TJ Lin, Gianni Carrozzi, Franco Barsanti, Alejandra Morales, Alison Mailing, Maria Espejo, Erica Gilbert, Louann Casano, Ellen O’Hara-Cicero, John Weed, Allen J. Dietrich

**Affiliations:** Clinical Directors Network (CDN); The Leukemia & Lymphoma Society; Montefiore Family Care Center; Urban Health Plan; Morris Heights Health Center; Lincoln Hospital Ambulatory Care Services; Segundo Ruiz Belvis Diagnostic &Treatment Center; Morrisania Diagnostic & Treatment Center; Good Shepherd Services; BronxWorks; Dartmouth College

**Keywords:** colorectal cancer screening, breast cancer screening, cervical cancer screening, depression, low-income women, care management, patient navigation, collaborative care, Federally Qualified Health Centers (FQHCs), Practice-based Research Network (PBRN)

## Abstract

**Background:** Women from low-income, racial/ethnic minority backgrounds receive fewer cancer screenings than other women *and* have higher rates of depression, which can interfere with cancer screening participation.

**Objective:** Assess the comparative effectiveness of two interventions to improve breast, cervical, and colorectal cancer screening and reduce depression among underserved women with depressive symptoms.

**Design:** Randomized comparative effectiveness trial

**Setting:** Six Federally Qualified Health Centers

**Participants:** N=757, 50-64 years with depression symptoms and overdue for cancer screening.

**Interventions:** Participants were randomized to collaborative depression care plus cancer screening intervention (*Collaborative Care Intervention*, CCI) or cancer screening intervention alone (*Prevention Care Management*, PCM). Both of these evidence-based, telephone interventions were delivered in English or Spanish, for up to 12 months, by care managers.

**Measurements:** Electronic Health Records provided cancer screening data (primary outcome). PHQ-9 measured depression at study entry (T0), 6- and 12-months post-baseline (T1 and T2, respectively; secondary outcome).

**Results:** Analyses revealed statistically significant increases in up-to-date status for all cancer screenings; depression improved in both intervention groups. There were no statistically significant differences between the two interventions in improving cancer screening rates or reducing depression.

**Limitations:** Depressive symptom improvement may be explained by typical symptom remission. The study duration (12-months) was insufficient to evaluate intervention effects on long-term cancer screening behavior and depression. This study was not powered for site-level analysis.

**Conclusions:** CCI and PCM both improved breast, cervical, and colorectal cancer screening and depression in clinical settings in underserved communities, however neither intervention showed an advantage in outcomes. Decisions about which approach to implement may depend on the nature of the practice and alignment of the interventions with other ongoing priorities and resources.

**Funding source:** This study was funded by Patient Centered Outcomes Research Institute (PCORI) (IH-12-11-4522) with additional infrastructure support from Agency for Healthcare Research and Quality (AHRQ) (5P30-HS-021667).

## Background and Objectives

Socioeconomic status is one of the strongest predictors of cancer screening behaviors (1-3) and severity (2, 4, 5). Low-income individuals are more likely to be diagnosed with late-stage cancers, due to lower rates of cancer screening, delayed follow-up of abnormal results (6), and less access to specialty care. Low-income women in the United States, and particularly racial/ethnic minority and low-income women, have fewer screenings for breast, cervical, and colorectal cancers and higher rates of mortality due to these cancers (6-10). In Bronx County, NY, the poorest urban county in the U.S., overall cancer deaths rates are 15% higher than New York City (11). The astonishing cervical cancer death rate is 60% higher than national rates (12), even though late-stage cervical cancer is largely preventable through screening (13).

In addition to these disparities in cancer screening, major depression is more common among low-income racial/ethnic minority women. Affecting roughly 20-25% of these women (14, 15), compared with ∼10% of US females (16), they suffer more chronic, severe, and disabling depression (17). Latinx and African American women are likely to seek support for depression in primary care settings (18). However, primary care depression management practices fall below evidence-based standards (19), despite the availability of effective treatments (20). Even when prompted by a primary care provider to seek mental health care, racial/ethnic minority women were less likely than white women to access specialty care and to take prescribed anti-depressant medication (21).

The observed disparities in both cancer screening and mental health care among low-income and racial/ethnic women have resulted in an alarming public health situation. There is growing evidence that women who experience untreated mental health problems (e.g., depression) are less likely to participate in cancer screenings (22, 23). Lower participation in cancer screening results in cancer diagnoses at later stages of tumor growth and metastasis, contributing to higher morbidity and mortality rates (6). Because this disparity widens among women from racial or ethnic minority groups (23), it is imperative to examine the intersectionality of sociodemographic factors on screening behaviors and depression treatment. That is, women who are both low-income and from racial/ethnic minorities may experience greater challenges related to cancer screening and depression treatment in additive and multiplicative ways, making them especially vulnerable to poor outcomes.

In the Bronx and nationally, cancer screening and mental health care needs have been identified as prime targets for improving patient health outcomes and well-being (24), yet no studies have attempted to intervene upon depression as a barrier to cancer screening. Clinical Directors Network’s (CDN) Prevention Care Management-3 (PCM3) study addressed this gap with a comparative effectiveness trial of “Collaborative Care Intervention (CCI)”, a novel, integrated intervention comprised of depression care management plus Prevention Care Management (PCM) versus PCM alone within Bronx Federally Qualified Health Centers (FQHCs). We hypothesized that CCI would be more effective than PCM for improving cancer screening participation for breast, cervical, and colorectal cancer (primary outcomes) and for reducing depression (secondary outcome) among low income women primarily of Latina, African American/Caribbean Black and other minority racial backgrounds.

## Methods

### Design Overview

This 1:1 randomized (CCI, PCM) comparative effectiveness trial was conducted in six Bronx, NY FQHCs. The study design, eligibility, and recruitment were previously described (25, 26). Because previous research showed that PCM is effective (27-29), ethics dictated offering an evidence-based intervention as the comparator. Study protocols were approved by sites’ Institutional Review Boards. Written informed consent was obtained upon enrollment.

### Setting and Participants

Eligible women were 50-64 years old, spoke English or Spanish, resided in Bronx, NY, screened positive for depression (Patient Health Questionnaire [PHQ]-9 ≥ 8), and were overdue for cancer screening according to US Preventive Services Task Force guidelines for at least one of the following: colorectal, breast, and/or cervical cancer (30-32).

Exclusion criteria included cancer diagnosis in the past six months, terminal illness, current pregnancy, or current suicidal thoughts. Women with suicidal thoughts were referred for care by their primary care provider. Care Managers reviewed Electronic Health Records to identify potentially eligible women; they were approached when they presented for care at their primary care provider appointment and were screened for depressive symptoms.

### Randomization and Interventions

#### Randomization

We used a randomized block algorithm, stratified by FQHC, to assign women to CCI or PCM. Block size was allowed to vary randomly to preclude any exercise of judgment or bias in the allocation of participants. The 1:1 allocation ratio was prepared by the offsite study statistician. The Data Coordinator maintained all randomization information offsite and only informed the enrolling Care Mangers of intervention assignments after baseline interviews were completed.

#### Intervention Components and Delivery

Table 1 compares components of the CCI and PCM interventions. A 12-month intervention period was chosen for each condition based on depression collaborative care (33-37) outcomes.

**Table 1.**
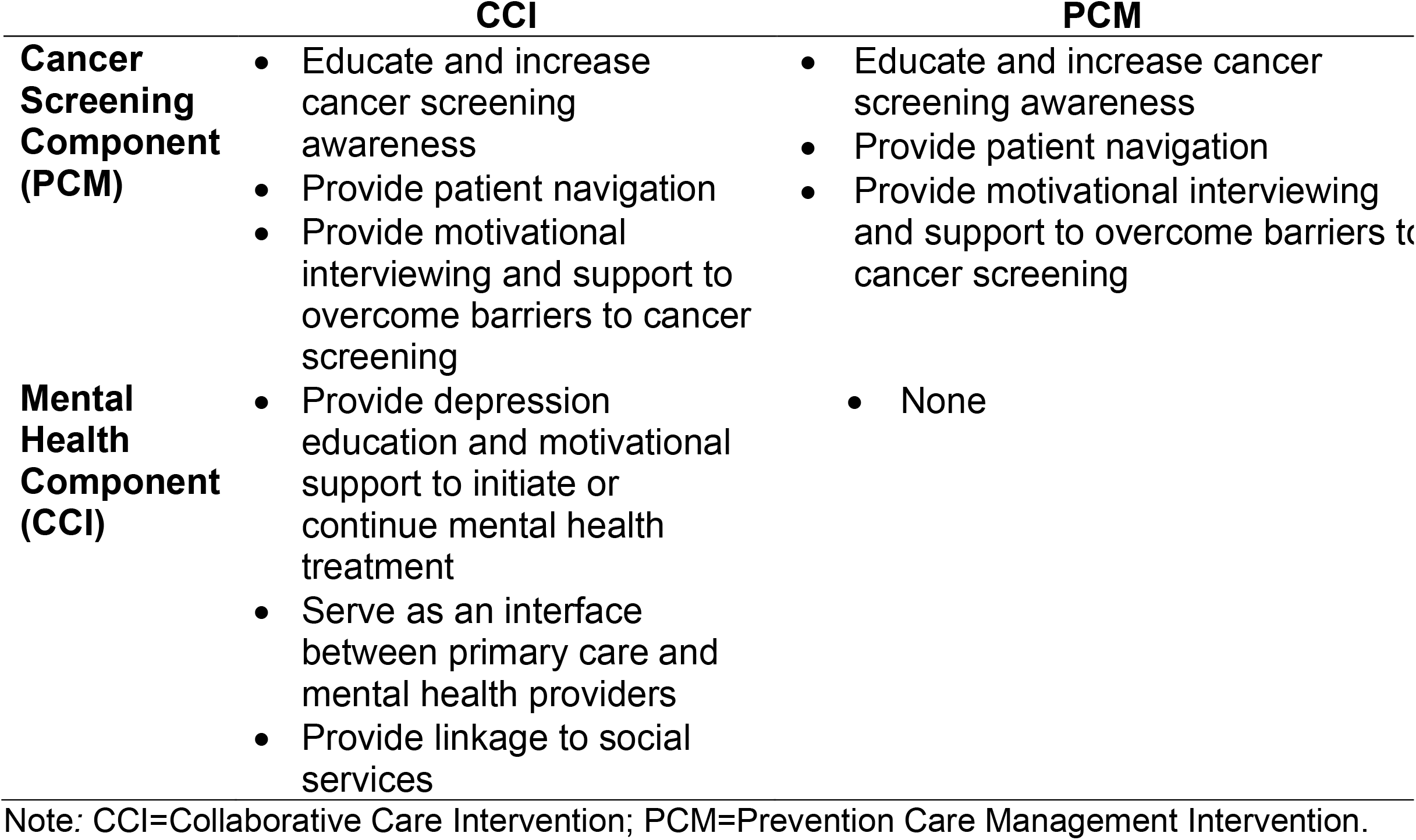
Intervention components

|  | CCI | PCM |
| --- | --- | --- |
| **Cancer Screening Component (PCM)** | • Educate and increase cancer screening awareness<br>• Provide patient navigation<br>• Provide motivational interviewing and support to overcome barriers to cancer screening | • Educate and increase cancer screening awareness<br>• Provide patient navigation<br>• Provide motivational interviewing and support to overcome barriers to cancer screening |
| **Mental Health Component (CCI)** | • Provide depression education and motivational support to initiate or continue mental health treatment<br>• Serve as an interface between primary care and mental health providers<br>• Provide linkage to social services | • None |

Note: CCI=Collaborative Care Intervention; PCM=Prevention Care Management Intervention.

##### Interventionists

Care Managers were female, bilingual (English-Spanish), college graduates, with experience working with underserved women. Participants always received the intervention from Care Managers located at their FQHC. Care Managers for both conditions performed similar patient navigation functions (e.g., scheduling primary care appointments, mailing educational materials, sending overdue cancer screening reminders). Care Managers were trained together in the PCM intervention; CCI interventionists underwent additional training. Each participating FQHC hired two separate Care Managers (one PCM, one CCI) to minimize the risk of contamination between the two study arms (38). We mitigated concerns about contamination between Care Managers at the same location who were delivering different interventions via extensive training (e.g., not sharing strategies and details of the study), supervision, and each Care Manager delivered only one intervention. We had no reports of information-sharing or intervention contamination among Care Managers.

##### Interventions

Both conditions were delivered by telephone to maximize time and efficiency and to enhance scale-up and sustainability, speculating that one care manager could work with patients across multiple sites in future studies. All study participants received English or Spanish language low health literacy educational materials about depression and cancer screening (39-42).

#### PCM

PCM is a telephone-based cancer screening support program shown to be effective in increasing rates of breast, cervical, and colorectal cancer screening across other trials in different kinds of primary care settings (28, 29, 43). Using a structured script, Care Managers informed participants of their group assignment, confirmed each participant’s cancer screening history, assessed readiness to engage in cancer screening, and collaborated with the participant to address cancer screening barriers (44). If a participant mentioned depressive symptoms, she was encouraged to make an appointment with her primary care clinician. PCM Care Managers contacted participants monthly until she became up-to-date for all three cancer screenings. When requested by the patient, Care Managers met in-person at the FQHCs for intervention visits. Participants who became up-to-date were sent a congratulatory card with due dates for upcoming cancer screenings.

#### CCI

In addition to the PCM activities, CCI Care Managers also evaluated participation in mental health treatment (past and present) and identified factors preventing adherence to mental health care recommendations. They also conducted motivational interviewing, encouraged mental health treatment, recommended mental health discussions with the primary care clinician, and connected women to social services at partnering community-based organizations. Care Managers documented the patient’s PHQ-9 depression score and their discussions on a feedback form provided to the primary care clinician (45). CCI Care Managers reached out by telephone at least once per month for up to 12 months. Care Managers conducted additional follow-up phone calls during the intervention period based on participants’ reported barriers and needs. Thus, while the intervention dose was not uniform, care was similar to real-world personalized patient-centered care.

### Outcomes and Follow-up

Assessments were conducted in English or Spanish at baseline (T0), 6 months (T1) and 12 months (T2) verbally over the telephone or in-person by trained project research assistants independent from Care Managers and blinded to treatment allocation. Participants were compensated a total of $40 for all completed assessments (45-60 minutes apiece).

#### Measures

##### Primary Outcome

Cancer Screening Behavior was the primary outcome, further divided into: breast, cervical, and colorectal cancer. Medical chart reviewers searched for: Pap testing (past 3 years without HPV; past 5 years with HPV), mammography (past 2 years), and colorectal screening (FOBT/FIT, in the past year; flexible sigmoidoscopy, in the past 5 years; and colonoscopy, in the past 10 years), and dichotomized as up-to-date (yes/no) using standardized items from the National Cancer Institute’s Health Information National Trends Survey (46). An ordinal composite variable was created reflecting the total number of up-to-date cancer screenings (0-3) at T0 and T2. Some participants may have received cancer screening outside the FQHCs, however, this information was routinely requested by practice staff for documentation.

##### Secondary Outcome

*Depression.* PHQ-9, a well-validated measure for screening and diagnosing depressive episodes, assessed depression severity and monitored treatment response (47-49). PHQ-9 is sensitive to depressive symptoms across racial and ethnic groups (47) and used in clinical practice. Measured at T0 and T2, respondents indicated how often they experienced depression symptoms in the past two weeks on a scale from 0 (not at all) to 3 (nearly every day). Responses were summed and higher scores indicated more severe depressive symptomatology. PHQ-9 ≥8 indicates clinically significant depressive symptoms (50). PHQ-9 scores were also subdivided into 0=no depression; 1-4=mild depression; 5-9=medium-mild; 10-14=moderate; 15-19=moderately severe; 20-27=severe depression (49).

##### Additional Secondary Outcomes

Secondary outcomes included patient-reported outcomes (listed in Appendix Table A1; collected at T0, T1, and T2).

### Statistical Analysis

#### Power and sample size

Sample size was calculated to detect a minimum 10% difference between arms in cancer screening participation with 80% power and 5% two-sided type I error rate. A 10% difference is considered the lowest difference of interest based on previous studies comparing PCM to usual care (27) to test the null hypothesis of no differences by condition; the alternative hypothesis was that the difference would be greater than 10% in cancer screening participation (with a two-sided Type I error rate of 5%). We estimated that 356 women in each arm was sufficient for 80% power to detect a 10% difference in screening participation between the two arms.

#### Analytic Plan

Analyses were conducted in SAS PROC GENMOD (V9.4). To describe baseline characteristics, continuous variables were summarized with means and standard deviations; group differences were analyzed with t-tests (e.g., number of calls completed during the trial) or repeated measures ANOVA. Categorical variables were analyzed with proportions or chi-square tests. Bivariate outcomes were analyzed using an unadjusted *χ*^2^ test separately for the three primary outcomes of cancer screening and *t*-test for the *a priori* secondary outcome of depression.

For the set of secondary analyses of cancer screening outcomes, multivariate logistic regression models examined study condition as a predictor of breast, cervical, and colorectal cancer screening behaviors at T2 in separate intention-to-treat models. Post-hoc multivariate logistic regression analyses included only women who were overdue for each cancer screening at T0. In addition, a proportional odds model was used to examine study condition as a predictor of total number of up-to-date cancer screening tests at T2 (range 0-3). All models included the following *a priori* covariates: age (years), household income (3 categories), T0 cancer screening status for the outcome of interest (up-to-date or not), and T0 depression (PHQ-9 score). FQHC site indicators were included in models as a fixed effect to account for clustering by site.

For depression, *a priori* secondary analyses used multivariate logistic regression to examine study condition as a predictor of PHQ-9 change. All models included *a priori* covariates: age (years), household income (3 categories), T0 cancer screening status (up-to-date or not), and T0 depression (PHQ-9 score). FQHC site indicators were included in the models as a fixed effect to account for clustering by site.

Further post-hoc analyses used separate multivariate logistic regressions to examine study condition as a predictor of (a) depression improvement and (b) depression remission from T0 to T2 controlling for T0 depression, number of up-to-date cancer screenings at T0, age, and household income. Depression improvement was defined as a reduction in severity grouping by at least one level (e.g., moderate depression at T0 and mild depression at T2). Depression remission was defined as a PHQ-9 score <5 at T2. The study was not powered for site-level analysis (see Appendix Table A2 for recruitment by FQHC site).

#### Missing data

Since missing data primarily resulted from missing PHQ-9s at T2 (N=129, 17% of participants), it was not appropriate to assume missing at random and to apply multiple imputation methods. Instead, we performed sensitivity analysis. In the model, those who were missing PHQ-9 at T2 were added as a separate category. We obtained similar intervention effects under both scenarios: OR=1.40 (1.02, 1.93), *p*=0.04 vs. OR=1.35 (0.98, 1.85) *p*=0.068.

## Results

### Patient Characteristics

In total, 802 participants were enrolled from 2014 to 2016 (Figure 1). Of these, 759 completed baseline interviews (95% response rate) and were randomized to either CCI (n=380) or PCM (n=379). Two participants were found to be ineligible after randomization because they were up-to-date for all three cancer screenings, resulting in 757 participants. Table 2 shows baseline demographic and clinical characteristics by study condition. There were no statistically significant demographic or clinical differences between study conditions at baseline (see also Appendix Table A3 for enrollment by racial categories and Table A4 for primary care utilization in the 18-months post-enrollment).

**Table 2.**
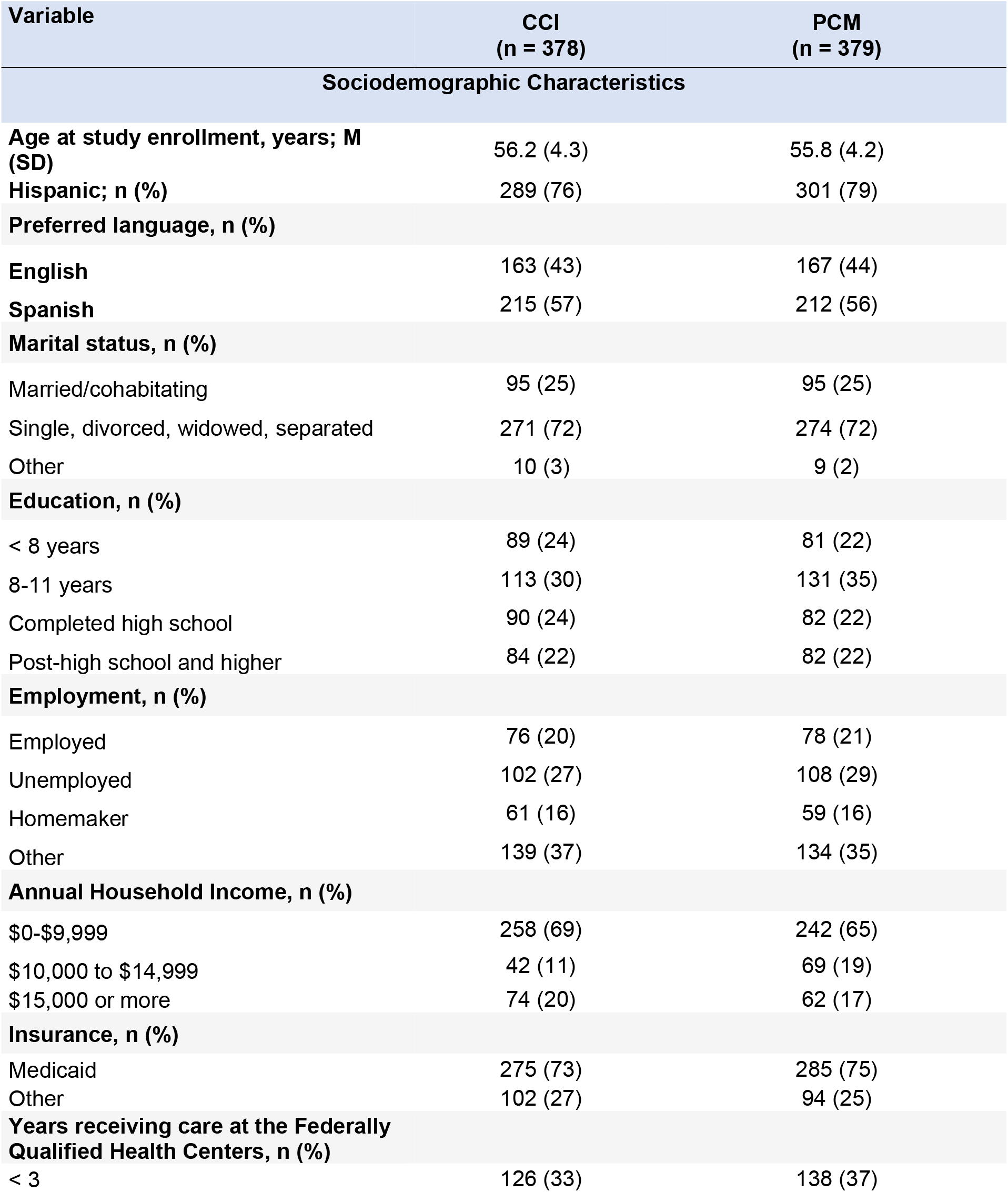
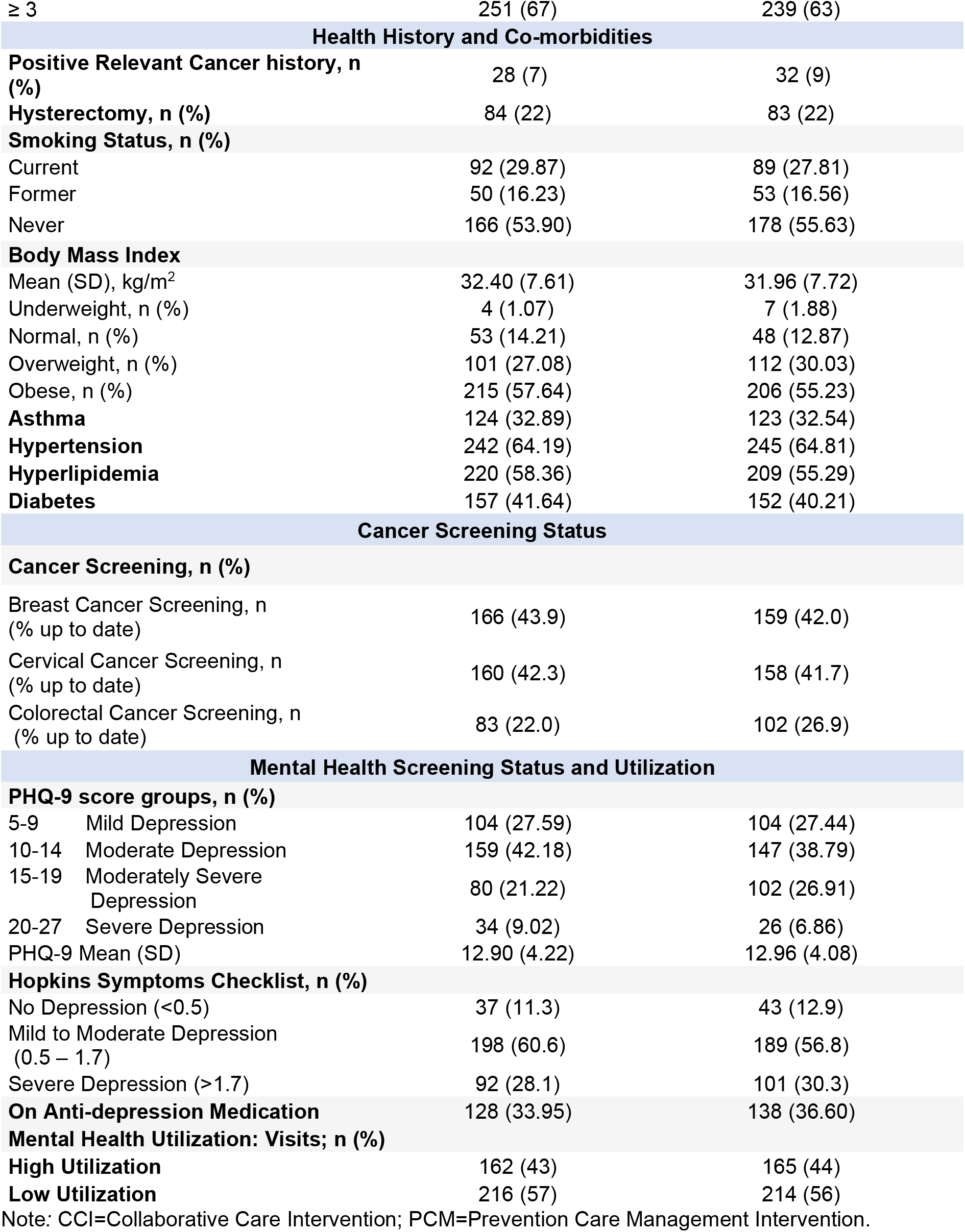
Participant baseline sociodemographic and clinical characteristics by study condition

| Variable | CCI<br>(n = 378) | PCM<br>(n = 379) |
| --- | --- | --- |
| **Sociodemographic Characteristics** |  |  |
| **Age at study enrollment, years; M (SD)** | 56.2 (4.3) | 55.8 (4.2) |
| **Hispanic; n (%)** | 289 (76) | 301 (79) |
| **Preferred language, n (%)** |  |  |
| English | 163 (43) | 167 (44) |
| Spanish | 215 (57) | 212 (56) |
| **Marital status, n (%)** |  |  |
| Married/cohabitating | 95 (25) | 95 (25) |
| Single, divorced, widowed, separated | 271 (72) | 274 (72) |
| Other | 10 (3) | 9 (2) |
| **Education, n (%)** |  |  |
| < 8 years | 89 (24) | 81 (22) |
| 8-11 years | 113 (30) | 131 (35) |
| Completed high school | 90 (24) | 82 (22) |
| Post-high school and higher | 84 (22) | 82 (22) |
| **Employment, n (%)** |  |  |
| Employed | 76 (20) | 78 (21) |
| Unemployed | 102 (27) | 108 (29) |
| Homemaker | 61 (16) | 59 (16) |
| Other | 139 (37) | 134 (35) |
| **Annual Household Income, n (%)** |  |  |
| \$0-\$9,999 | 258 (69) | 242 (65) |
| \$10,000 to \$14,999 | 42 (11) | 69 (19) |
| \$15,000 or more | 74 (20) | 62 (17) |
| **Insurance, n (%)** |  |  |
| Medicaid | 275 (73) | 285 (75) |
| Other | 102 (27) | 94 (25) |
| **Years receiving care at the Federally Qualified Health Centers, n (%)** |  |  |
| < 3 | 126 (33) | 138 (37) |
| ≥ 3 | 251 (67) | 239 (63) |
| **Health History and Co-morbidities** |  |  |
| **Positive Relevant Cancer history, n (%)** | 28 (7) | 32 (9) |
| **Hysterectomy, n (%)** | 84 (22) | 83 (22) |
| **Smoking Status, n (%)** |  |  |
| Current | 92 (29.87) | 89 (27.81) |
| Former | 50 (16.23) | 53 (16.56) |
| Never | 166 (53.90) | 178 (55.63) |
| **Body Mass Index** |  |  |
| Mean (SD), kg/m<sup>2</sup> | 32.40 (7.61) | 31.96 (7.72) |
| Underweight, n (%) | 4 (1.07) | 7 (1.88) |
| Normal, n (%) | 53 (14.21) | 48 (12.87) |
| Overweight, n (%) | 101 (27.08) | 112 (30.03) |
| Obese, n (%) | 215 (57.64) | 206 (55.23) |
| **Asthma** | 124 (32.89) | 123 (32.54) |
| **Hypertension** | 242 (64.19) | 245 (64.81) |
| **Hyperlipidemia** | 220 (58.36) | 209 (55.29) |
| **Diabetes** | 157 (41.64) | 152 (40.21) |
| **Cancer Screening Status** |  |  |
| **Cancer Screening, n (%)** |  |  |
| Breast Cancer Screening, n (% up to date) | 166 (43.9) | 159 (42.0) |
| Cervical Cancer Screening, n (% up to date) | 160 (42.3) | 158 (41.7) |
| Colorectal Cancer Screening, n (% up to date) | 83 (22.0) | 102 (26.9) |
| **Mental Health Screening Status and Utilization** |  |  |
| **PHQ-9 score groups, n (%)** |  |  |
| 5-9 Mild Depression | 104 (27.59) | 104 (27.44) |
| 10-14 Moderate Depression | 159 (42.18) | 147 (38.79) |
| 15-19 Moderately Severe Depression | 80 (21.22) | 102 (26.91) |
| 20-27 Severe Depression | 34 (9.02) | 26 (6.86) |
| PHQ-9 Mean (SD) | 12.90 (4.22) | 12.96 (4.08) |
| **Hopkins Symptoms Checklist, n (%)** |  |  |
| No Depression (<0.5) | 37 (11.3) | 43 (12.9) |
| Mild to Moderate Depression (0.5 – 1.7) | 198 (60.6) | 189 (56.8) |
| Severe Depression (>1.7) | 92 (28.1) | 101 (30.3) |
| **On Anti-depression Medication** | 128 (33.95) | 138 (36.60) |
| **Mental Health Utilization: Visits; n (%)** |  |  |
| **High Utilization** | 162 (43) | 165 (44) |
| **Low Utilization** | 216 (57) | 214 (56) |

Note: CCI=Collaborative Care Intervention; PCM=Prevention Care Management Intervention.

**Figure 1.**
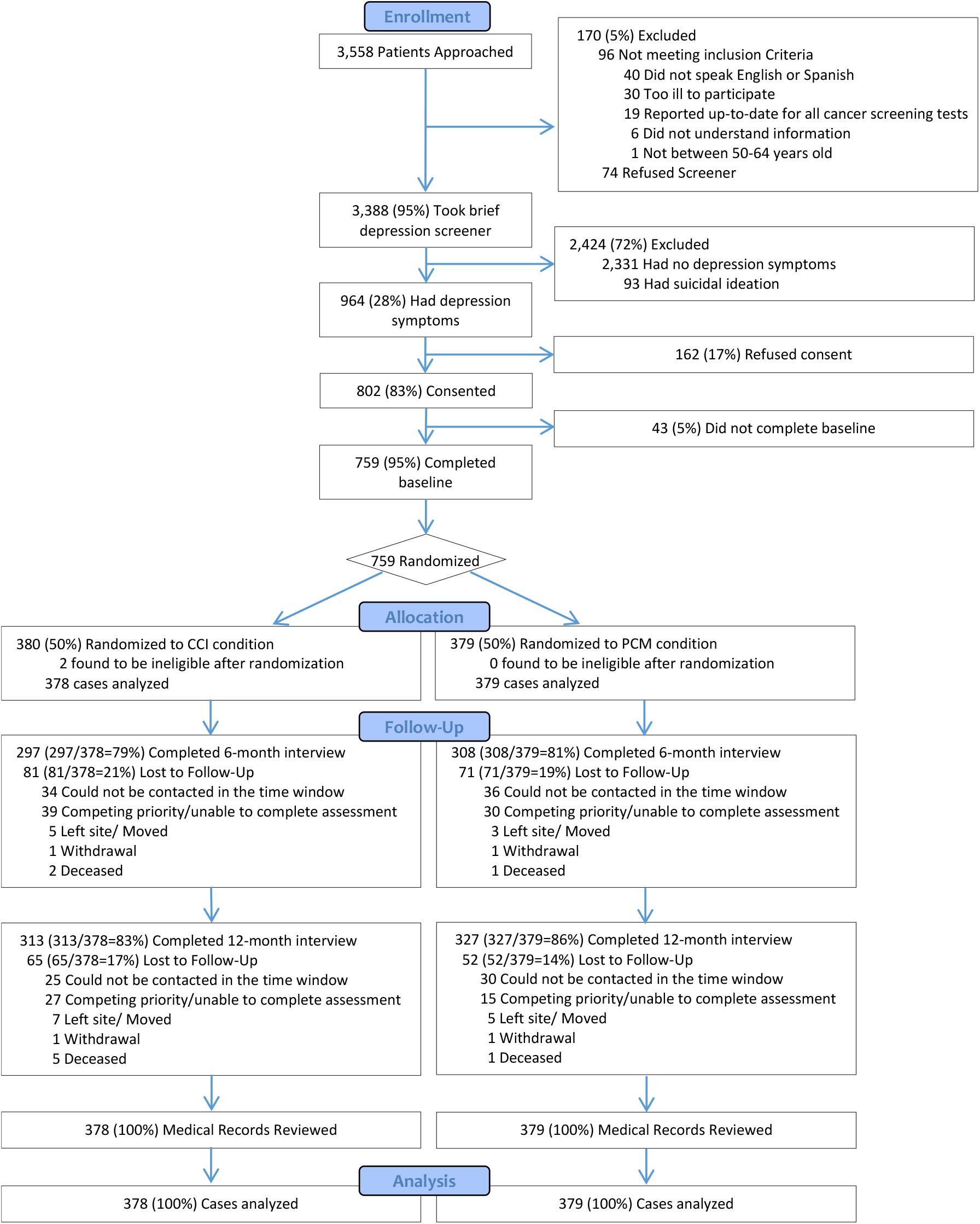
CONSORT flow diagram

### Intervention Delivery

Most interactions occurred by phone (90%). On average, women in the CCI arm completed six calls (range 0-20), while women in the PCM arm completed three (range 0-17; *p*<0.001). Initial calls for CCI averaged 42 minutes (range 4-118 minutes) compared to 13 minutes (range 4-82 minutes) for PCM. Subsequent calls averaged 18 minutes for CCI (range 2-101 minutes) and 9 minutes (range 1-62 minutes) for PCM, providing evidence of treatment fidelity.

### Differences among conditions at baseline and 12-month follow-up

For breast cancer screening, we found no differences for CCI vs. PCM at baseline (43.9% vs. 42.0%) or at follow up (66.7% vs. 66.8%) (intervention arm differences *p*=.98). For cervical cancer screening, we found no differences for CCI vs. PCM at baseline (42.3% vs. 41.7%) or at follow up (61.6% vs. 64.6%) (intervention arm differences *p*=.39). For colorectal cancer screening, we found no differences for CCI vs. PCM at baseline (22.0% vs. 26.9%) or at follow up (56.9% vs. 52.8%) (intervention arm differences *p*=.26).

### Primary Outcomes

#### Breast Cancer Screening

There was no statistically significant difference between the two intervention arms for improving breast cancer screening participation over time. From baseline to follow-up, the proportion of participants up-to-date for breast cancer screening increased significantly in both arms (Table 3; CCI: 43.9% to 66.7% and PCM: 42.0% to 66.8%).

**Table 3.** Proportion of participants who were up-to-date for cancer screening from study entry to 12-month follow-up

|  | **CCI**<br>**(n= 378)** | **PCM**<br>**(n= 379)** | **Chi-square for time**<br>**point** | **p-value for**<br>**timepoint** |
| --- | --- | --- | --- | --- |
| **Breast**<br>**Cancer** |  |  |  |  |
| Baseline | 43.9% | 42.0% | 0.30 | .59 |
| Follow-up | 66.7% | 66.8% | 0.0007 | .98 |
| **Cervical**<br>**Cancer** |  |  |  |  |
| Baseline | 42.3% | 41.7% | 0.03 | .86 |
| Follow-up | 61.6% | 64.6% | 0.73 | .39 |
| **Colorectal**<br>**Cancer** |  |  |  |  |
| Baseline | 22.0% | 26.9% | 2.52 | .11 |
| Follow-up | 56.9% | 52.8% | 1.29 | .26 |

Note: For chi-square analyses, all degrees of freedom = 1, sample size = 757.
CCI=Collaborative Care Intervention; PCM=Prevention Care Management Intervention.

In an *a priori* logistic regression model of only women who were overdue for breast cancer screening at T0, only lower depression at T0 significantly predicted being up-to-date for breast cancer screening at T2 (*p*=0.048) (Table A6). As up-to-date at baseline may be a non-intuitive control, we calculated a 2×2 by timepoint up-to-date matrix; 33% of women who were up-to-date for breast cancer screening at baseline were also up-to-date at T2.

### Cervical Cancer Screening

There was no statistically significant difference between the two arms for improving cervical cancer screening participation over time. From T0-T2, the proportion up-to-date increased significantly in both study arms (Table 3; CCI: 42.3% to 61.6% and PCM: 41.7% to 64.6%).

In an *a priori* analysis of only women who were overdue for cervical cancer screening at T0, only lower depression at T0 significantly predicted being up-to-date for cervical cancer screening at T2 (*p*=0.03) (Table A6). In the 2×2 up-to-date by timepoint matrix; 63% of women who were up-to-date for cervical cancer screening at baseline were also up-to-date at T2.

### Colorectal Cancer Screening

There was no statistically significant difference between the intervention arms for improving colorectal cancer screening status. The proportion up-to-date increased significantly in both study arms from baseline to T2 (Table 3; CCI: 22.0% to 56.9% and PCM: 26.9% to 52.8%).

In an *a priori* subgroup analysis of women who were overdue for colorectal cancer screening at T0, there was a significant treatment effect on screening status at T2 (Table A6). Specifically, among women who were overdue for colorectal cancer screening at T0, women randomized to CCI were more likely to be up-to-date for colorectal cancer screening at T2 compared to women randomized to PCM (OR=1.57; 95% CI 1.04-2.37) (*p*=0.030). Younger age and lower income also significantly predicted being up-to-date at T2 in this model. In the 2×2 up- to-date by timepoint matrix; 24% of women who were up-to-date for colorectal cancer screening at baseline were also up-to-date at T2.

### Secondary Outcomes

#### Depression

##### Continuous depression scores

From baseline to follow-up, depression scores significantly decreased in both arms (Table 4). PHQ-9 scores significantly declined from baseline to 6-m follow-up *and* baseline to 12-m follow-up. There was no statistically significant difference between the two arms in mean depressive symptoms over time. For mental health visits, there were no differences by condition at baseline (*p*=0.85), T1 (*p*=0.23), or T2 (*p*=0.98). For prescription medications for depression, there were no differences between conditions at T0 (*p*=0.57), T1 (*p*=0.92), or T2 (*p*=0.99).

**Table 4.** Change over time in depression scores from study entry to 12-month follow-up

| Condition | Baseline Mean | Six-Month Follow-up Mean | 12-month Follow-up Mean | t-statistic baseline -6m | p value baseline -6m | 95% Confidence Interval | t-statistic baseline -12m | p value | 95% Confidence Interval |
| --- | --- | --- | --- | --- | --- | --- | --- | --- | --- |
| CCI | 12.86 | 7.88 | 8.23 | 13.70 | .000 | 3.88-5.17 | 12.04 | .000 | 3.84-5.35 |
| PCM | 12.68 | 8.34 | 7.84 | 13.24 | .000 | 4.09-5.51 | 14.08 | .000 | 4.34-5.76 |

Note: CCI=Collaborative Care Intervention; PCM=Prevention Care Management Intervention. Depression was measured by PHQ-9 continuous scores. In a post-hoc analysis (2 (condition) x 3 (time point: baseline, 6-month, 12-month) repeated measures ANOVA, there were no group by time effects ( $F(2, 536) < 1$ ) indicating that both conditions saw statistically significant decreases over the course of the study but the conditions showed indistinguishable patterns when compared to each other.

Additional secondary outcome analyses comparing CCI and PCM at different time points are presented in Appendix Table A5.

In heterogeneity of treatment effect analyses using primary language, the odds ratio for breast cancer screening was statistically significant, whereas cervical and colorectal cancer screening were not (Table 5).

**Table 5.**
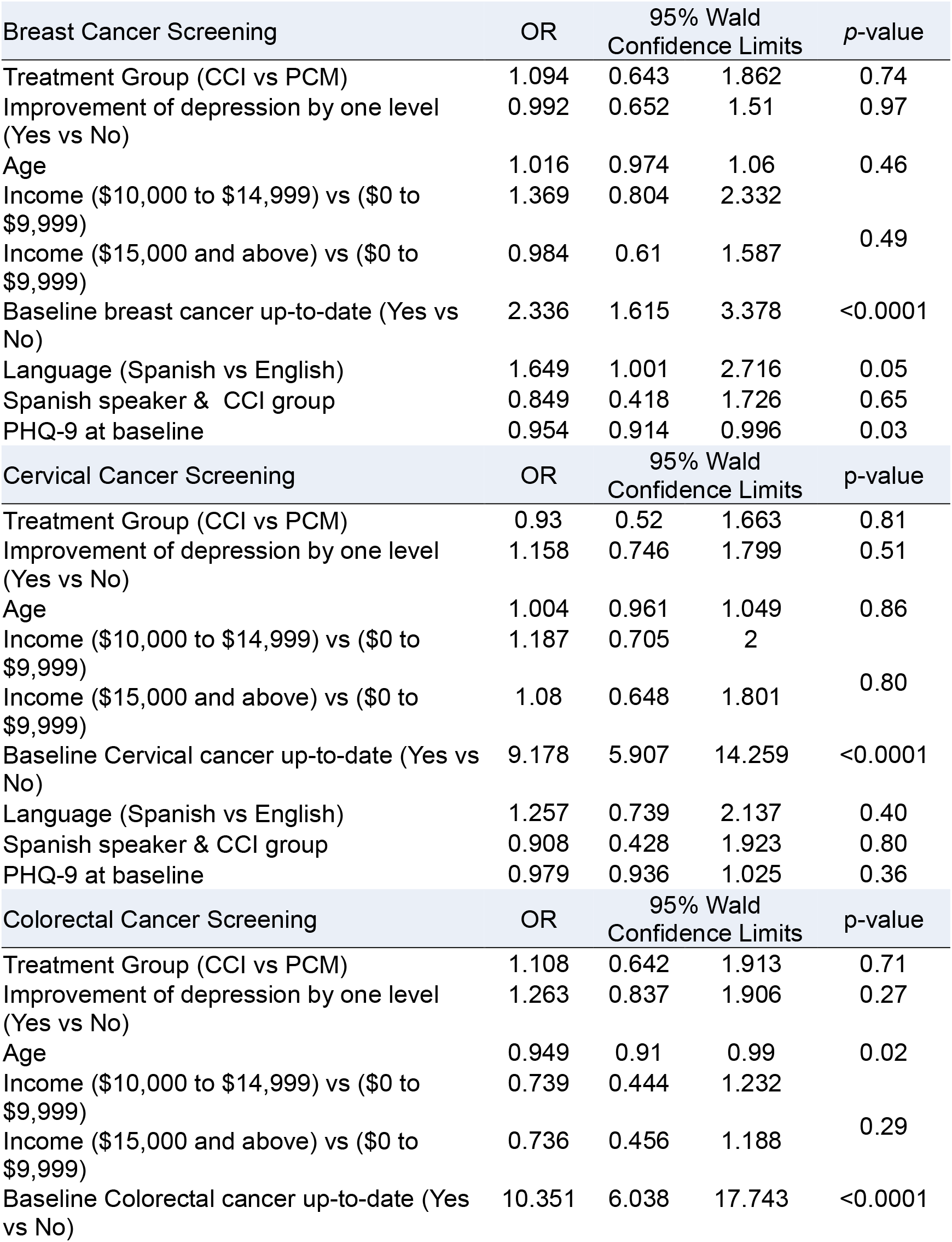
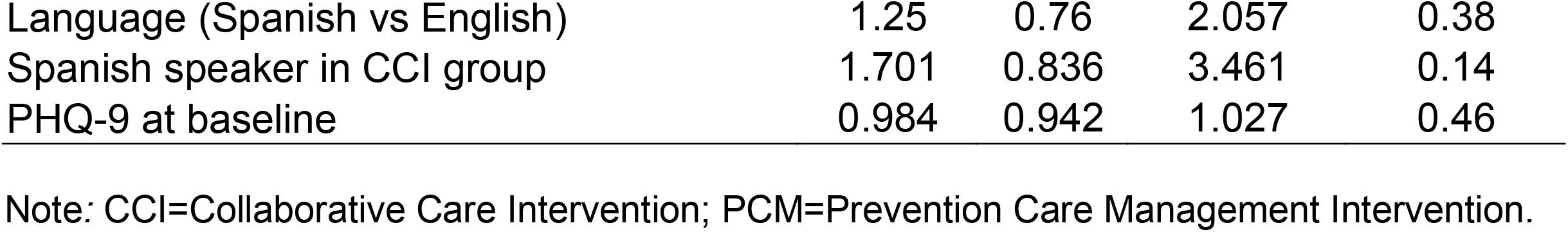
Heterogeneity analysis on primary language for breast cancer screening, cervical cancer screening, and colorectal cancer screening

### Exploratory Analyses

#### Overall Cancer Screening

There was no statistically significant difference between the two intervention arms for improving total number of up-to-date cancer screenings from T0 to T2 (Table A7). Being up-to-date for one cancer screening at T0 significantly predicted the achievement of additional up-to-date screenings at T2. This relationship was stronger for women who were up-to-date for two cancer screenings at T0.

#### Depression improvement and remission

Comparing the subset of women who were followed vs. not followed on depression measures, ethnicity emerged as the only group difference (Table A8). The intervention arms did not differentially improve depression by ≥1 severity level (Table A9). Greater depression at T0 was the only significant predictor of level of depression improvement from T0-T2 (*p*=0.002). Similarly, the conditions did not differ for depression remission (Table A9). Household income (*p*=0.037) and lower depression (*p*<0.001) at T0 were significant predictors of depression remission from T0-T2.

## Discussion

The PCM3 study was a randomized clinical trial comparing the effectiveness of two evidence-based care management interventions to improve cancer screening among underserved women with depression in Bronx, NY. Both CCI and PCM produced clinically meaningful and statistically significant increases in breast, cervical, and colorectal cancer screening rates, as well as improvements in depressive symptoms from baseline to 12-month follow-up. However, we did not observe any statistically significant differences between the two treatment arms for the primary outcomes (breast, cervical and colorectal cancer screening) and secondary outcome (change in depression).

When examining *a priori* analyses of women who were overdue for breast cancer and cervical cancer screening at baseline, we observed that lower depression at baseline was the only predictor of being up-to-date at follow-up. For participants who were overdue for colorectal cancer screening at baseline, women randomized to CCI were more likely to be up-to-date for colorectal cancer screening at T2 compared to women randomized to PCM. Younger age and higher income were also significant predictors of up-to-date colorectal screening status at follow-up. Exploratory analyses revealed no differential treatment effects for overall cancer screening for the three types of cancer combined, depression improvement, or depression remission.

As a comparison to the published literature (using “interventions in primary care settings” as the English-language search term), PCM was shown to be effective in increasing cancer screening (27-29, 45, 51) and CCI was shown to reduce symptoms of depression (40, 48). In this comparative effectiveness trial, however, the main finding was that neither intervention proved to be more effective than the other. Women benefitted from care management that included patient navigation and support to undergo cancer screenings, regardless of whether depression was also targeted during these interactions. It is possible that we did not find differential treatment effects between CCI and PCM because the patient navigation support provided by the Care Managers across both conditions strengthened the relationships between patients and their care providers, and both approaches emphasized patient activation and self-management. Given the greater investment of time and resources needed to implement the CCI intervention (e.g., training in the PCM intervention alone requires less time, and an average of six longer calls for CCI versus three shorter calls for PCM), it may be more appealing to payers, health centers and other stakeholders to implement PCM.

With regard to generalizability and limitations, the lack of differential intervention effects on depression-related outcomes may have resulted from a combination of the natural course of depression remission plus access to co-located/integrated primary care and mental health services in participating FQHCs. Larger intervention effects might have been evident in the CCI arm if participants did not have access to onsite mental health services. While it may be difficult to justify the addition of depression care management to the PCM intervention at practices with onsite mental health care, CCI might confer greater benefits for cancer screening and depression outcomes in clinical settings that do not have integrated mental health services.

Participants were required to have elevated depression scores upon enrollment, but we did not assess perceptions of unmet mental health care needs. Future work should consider enrolling patients who demonstrate unmet mental health care needs or some level of underutilization of care. While recruitment would be slower, this might provide a better test of the CCI intervention.

This study was conducted over a 12-month period with assessments at 6- and 12-months post-baseline. Perhaps differential intervention effects would have emerged with a longer follow-up, especially given the varying time frames for screening recommendations, ranging from every two years for mammograms to every 10 years for colonoscopy. Future work could also include additional patient-reported measures, such as behavioral activation.

Sociodemographic factors might affect the uptake of cancer screening guidelines. Though our analyses were limited to data already collected, we conducted additional subgroup heterogeneity analyses on primary language (Table 5). While not all Latinas spoke Spanish, all Spanish-speakers identified as Latina. With language in the model, the OR was 1.65 for breast cancer screening; cervical and colorectal cancer screening were non-significant. We did not have other adequately powered subgroups to make meaningful group comparisons. Future studies should consider stratifying and oversampling in order to investigate heterogeneity of treatment effects within other subgroups.

This trial demonstrated that CCI and PCM can be implemented successfully in medically underserved communities and is generalizable to diverse populations. Both interventions offer strong potential for implementation and scalability. They are protocol-driven, evidence-based interventions that were designed to be delivered by health care staff (e.g., health educators, care coordinators, patient navigators). Manuals and formalized training provide the tools, barriers-based scripts, and detailed procedures for implementation in both English and Spanish (41).

In conclusion, CCI and PCM interventions were both similarly effective in increasing cancer screening and reducing depression among underserved women who were from primarily Latina, African American/Caribbean Black and other racial minority backgrounds. CCI and PCM interventions are implementable, scalable, effective programs which will enable health care systems to detect cancers at an earlier stage to prevent excess morbidity and mortality associated with breast, cervical, and colorectal cancer among underserved women with depression. As we did not observe superiority of one intervention over the other, it may be more appealing to health providers to implement PCM to enhance cancer screening among underserved women.

## Data Availability

The Study Protocol (original and amendments), computer code that authors used to generate the results reported in the published article, and the documented analytic dataset will be available to approved individuals through written agreements with the lead author. 
Protocol: https://www.pcori.org/sites/default/files/Tobin229-Final-Research-Report.pdf.
Statistical Code: Available to interested readers by contacting Dr. Tobin at.
Data: Available to interested readers by contacting Dr. Tobin at.

## Acknowledgements

We wish to acknowledge the following practices and Community Based Organizations (CBOs) for their participation: Montefiore Family Care Center, Morris Heights Health Center, NYC Health+Hospitals (Gotham Health/Morrissania, Gotham Health/Segundo Ruiz Belvis, Lincoln Ambulatory Care Services), Urban Health Plan, BronxWorks, and Good Shepherd Services.

We would like to acknowledge the project team members and stakeholders from the participating sites and CBOs: Norma Perez, Anitta Ruiz, Edgar Gomez, MD, and Maryanne Guerrero, MPH at NYC Health+Hospitals (Gotham Health/Morrissania, Gotham Health/Segundo Ruiz Belvis, Lincoln Ambulatory Care Services).

---

We also wish to thank the research team members from Clinical Directors Network: Carmen Rodriguez, Cleo Clarize Overa, Elia Flores, Genevieve Lalanne-Raymond, RN, MPH, Jackie Cortez, Maria Sae-Hau, PhD, Melissa Coronado, Niurka Vidal, Sandra Monroy, Sarah Wagoner, Sarah Johnson, MD, MPH, as well as Suzanne Lechner, Ph.D. for editorial assistance, and Dr. Ardis Olson for review of the manuscript.

Current Full Mailing Addresses for All Authors

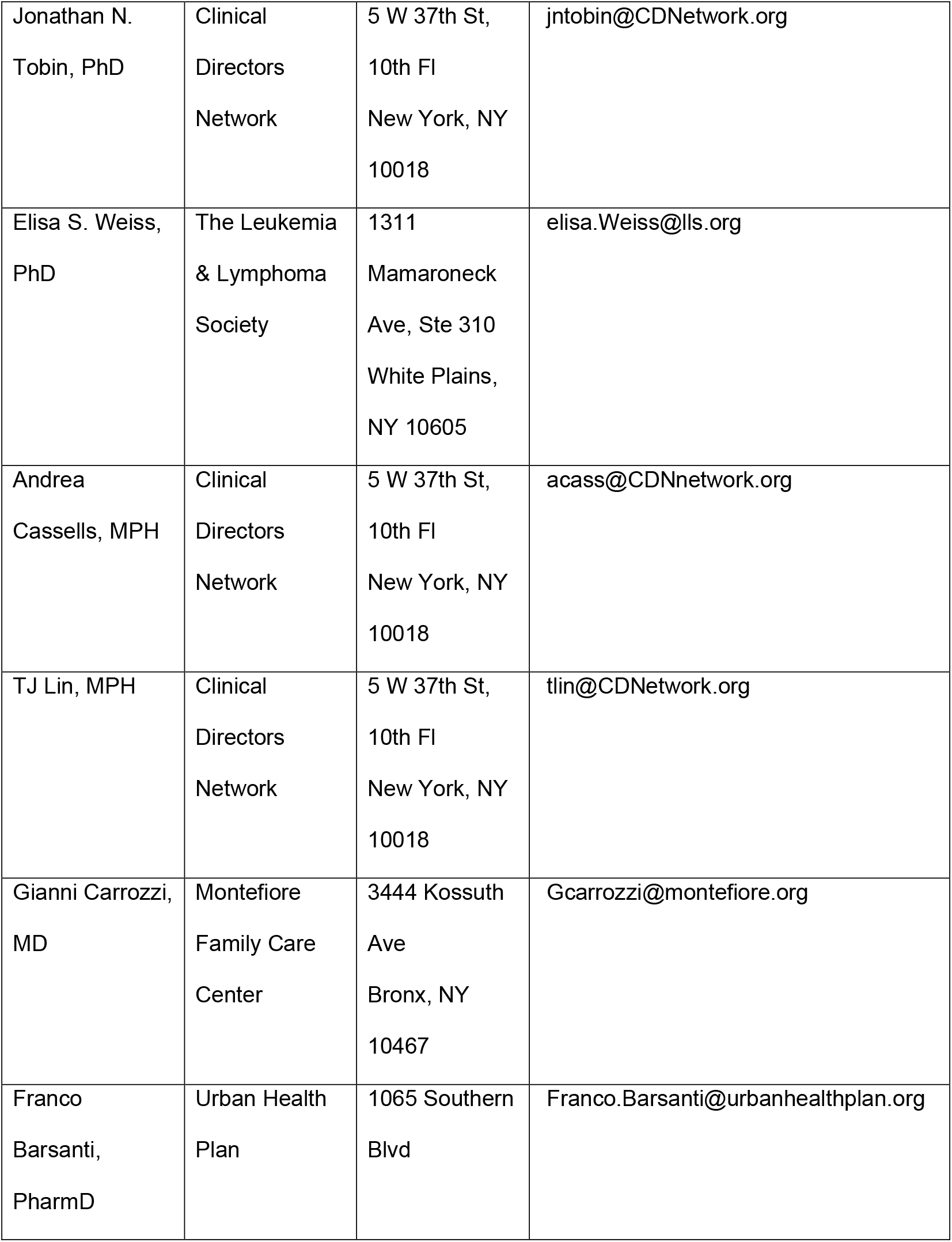
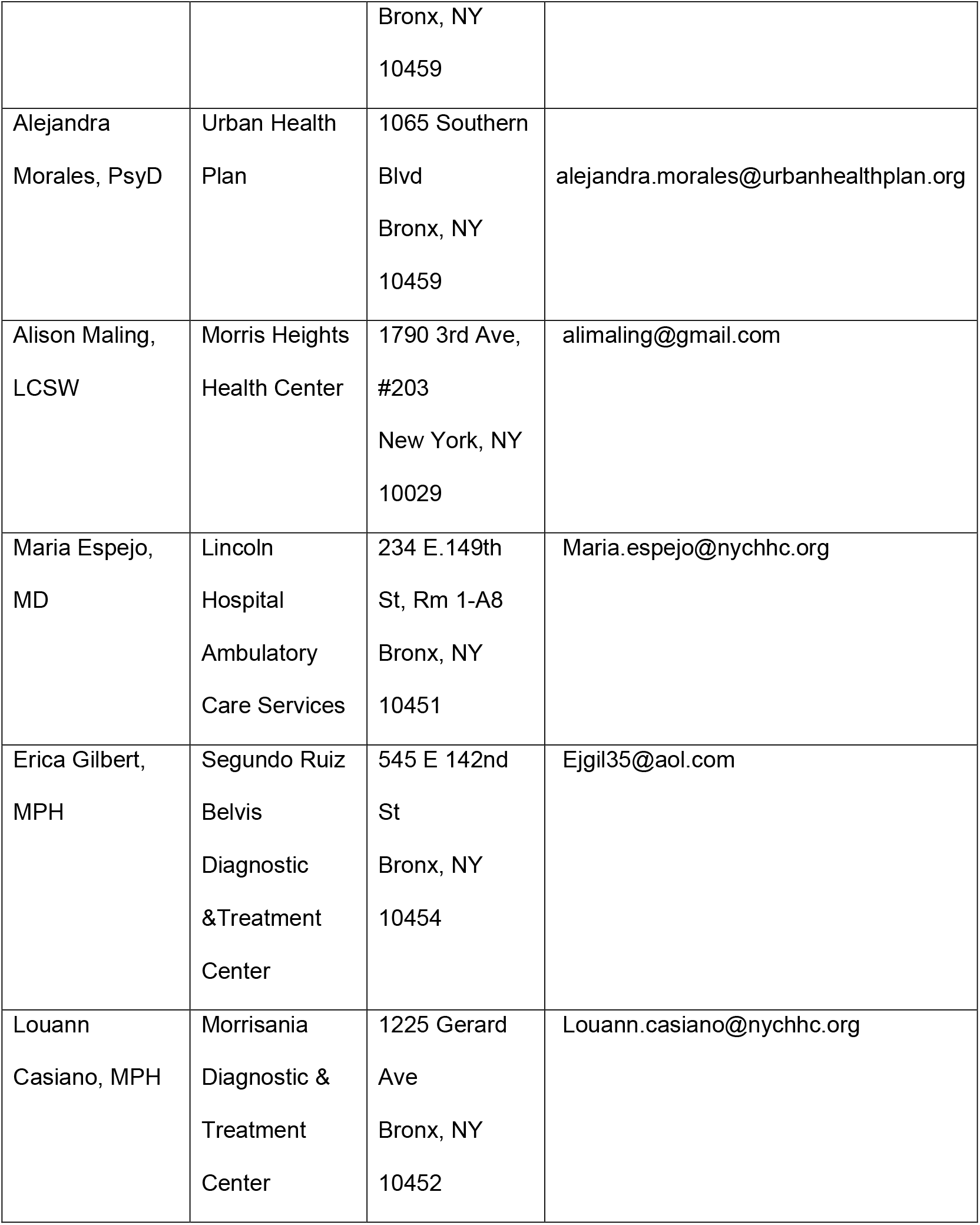
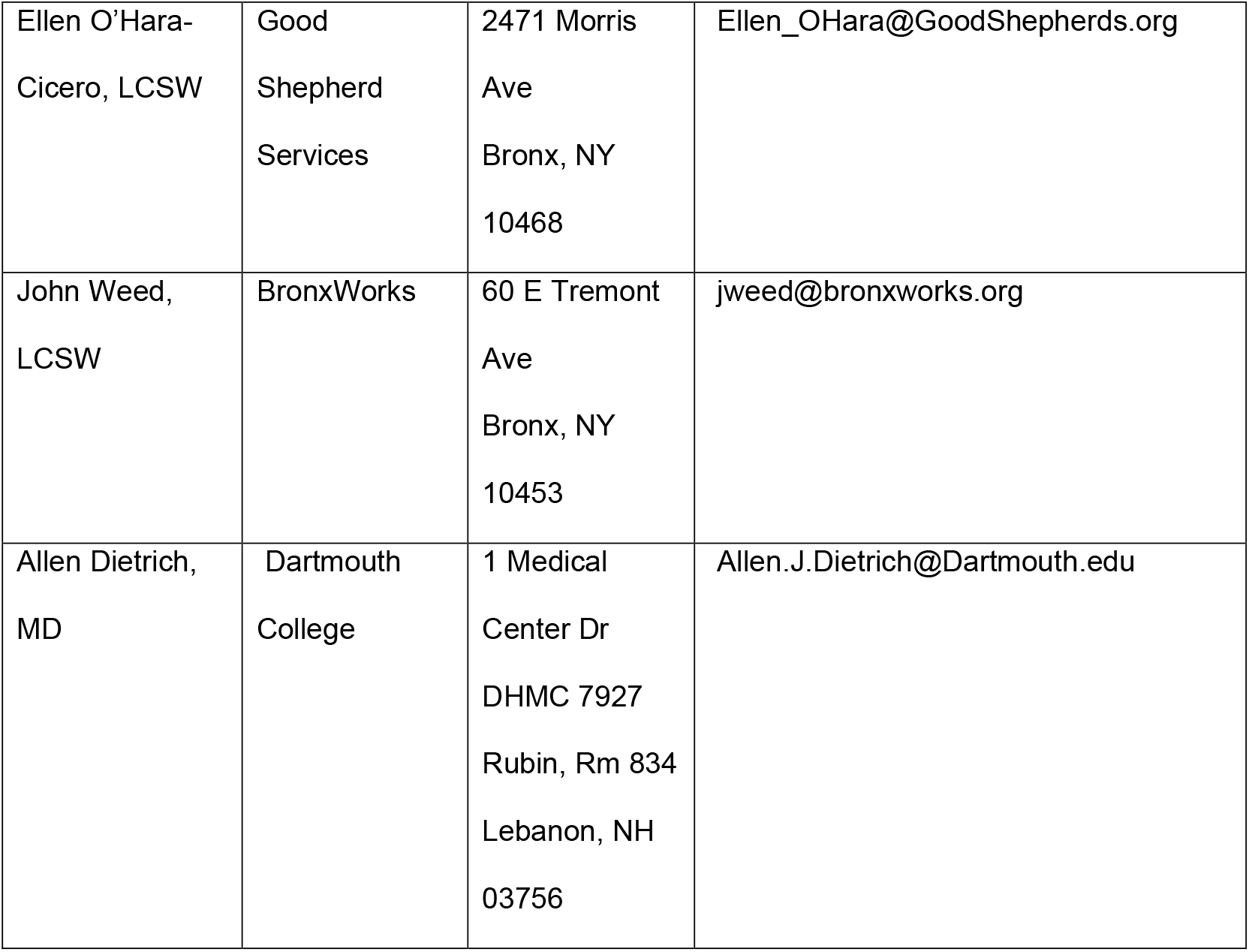

**APPENDIX Table A1.**
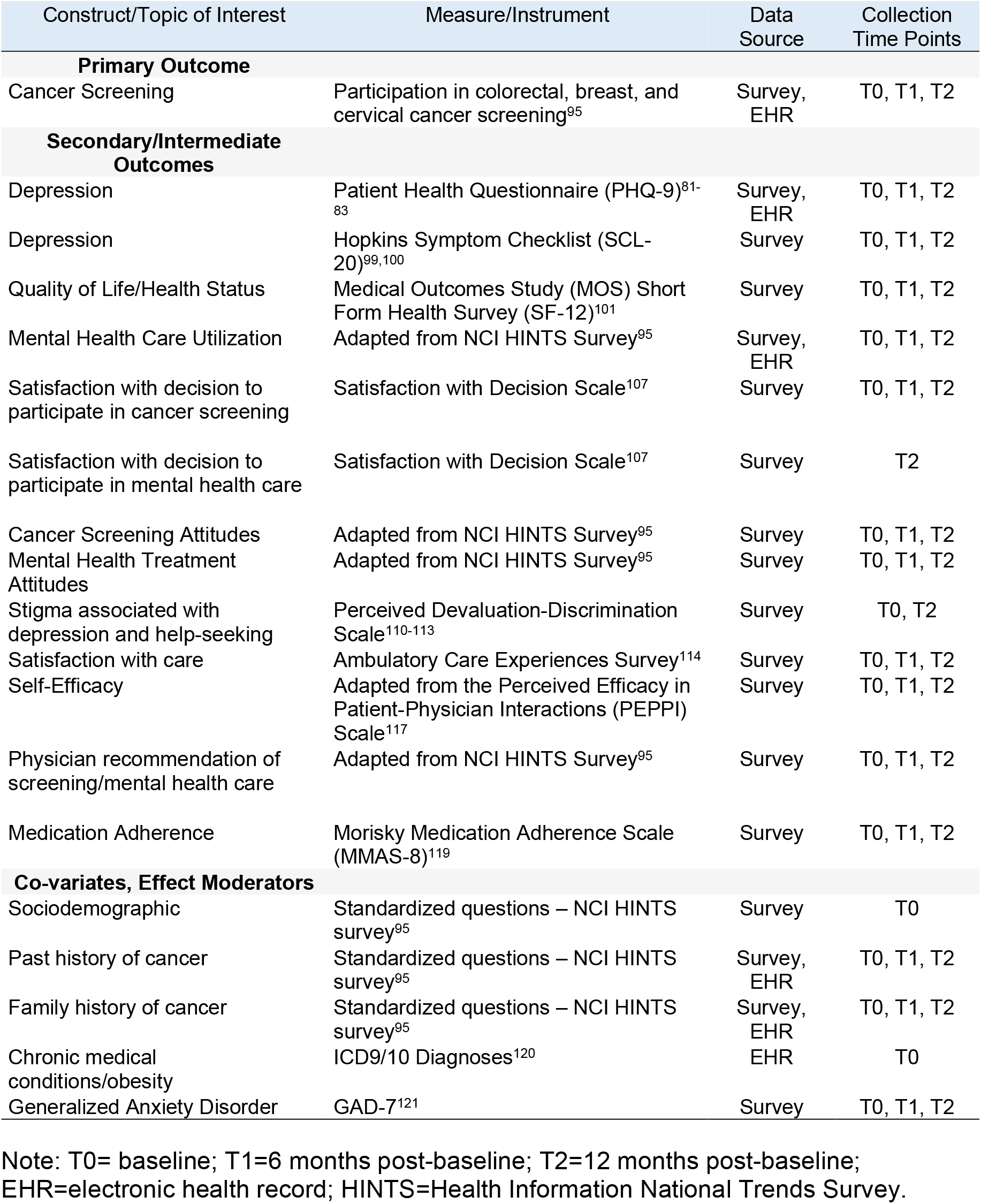
Study measures, data source, and timing

**APPENDIX Table A2.**
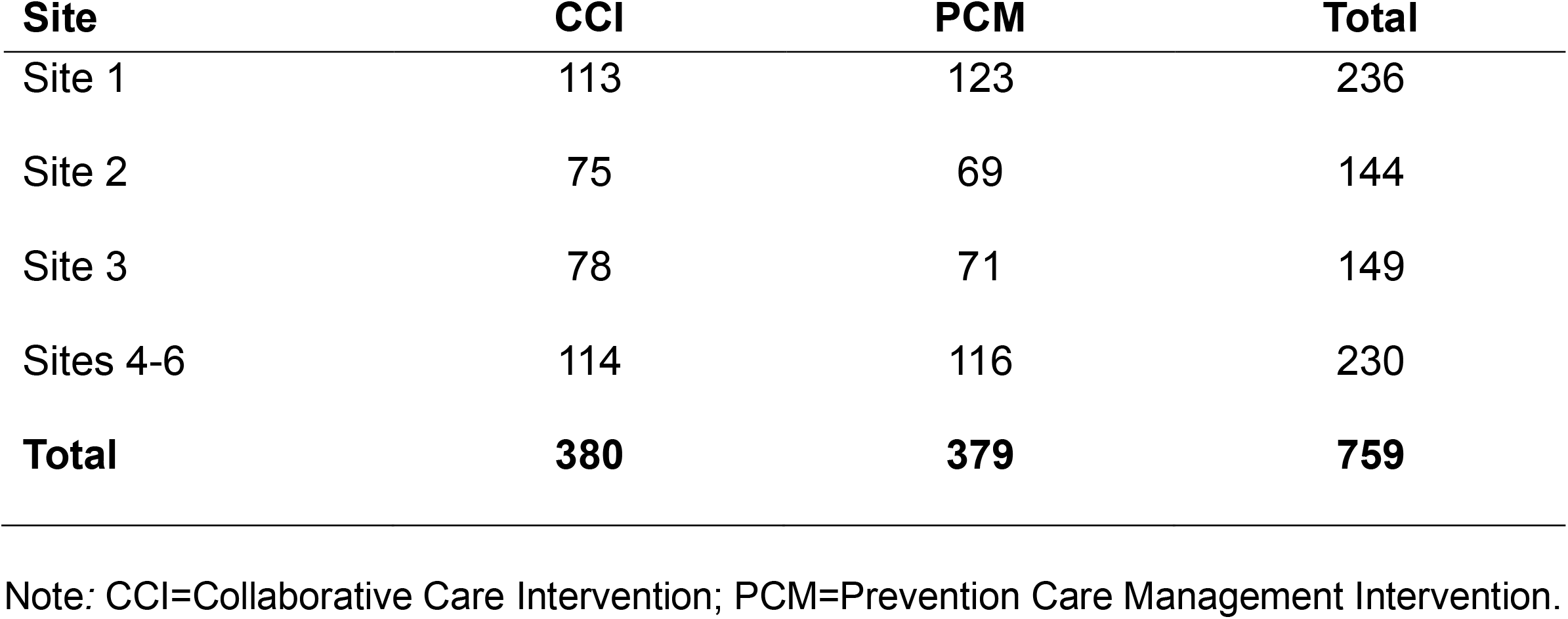
Patient recruitment by Federally Qualified Health Center Site

| **Site** | **CCI** | **PCM** | **Total** |
| --- | --- | --- | --- |
| Site 1 | 113 | 123 | 236 |
| Site 2 | 75 | 69 | 144 |
| Site 3 | 78 | 71 | 149 |
| Sites 4-6 | 114 | 116 | 230 |
| **Total** | **380** | **379** | **759** |

Note: CCI=Collaborative Care Intervention; PCM=Prevention Care Management Intervention.

**APPENDIX Table A3.** Racial enrollment table

| Self-Report of Race | N |
| --- | --- |
| American Indian/Alaska Native | 5 |
| Asian | 0 |
| Black/African American | 157 |
| Hawaiian/Pacific Islander | 0 |
| White | 25 |
| Multi-race | 36 |
| Other | 579 |

Note: Self-reported race based on US Census Bureau categories. Most women of Latina/Hispanic background endorsed “Other” or “White” categories.

**APPENDIX Table A4.** Primary care utilization in 18-months post consent

|  | CCI (N= 378) | PCM (N= 379) |
| --- | --- | --- |
| 0 visit | 12 (3.2%) | 7 (1.8%) |
| 1-3 visits | 65 (17.2%) | 63 (16.6%) |
| 4-6 visits | 87 (23.0%) | 90 (23.7%) |
| 7-9 visits | 103 (27.2%) | 86 (22.7%) |
| 10 and more visits | 111 (29.4%) | 133 (35.1%) |

Note: CCI=Collaborative Care Intervention; PCM=Prevention Care Management Intervention.

**APPENDIX Table A5.**
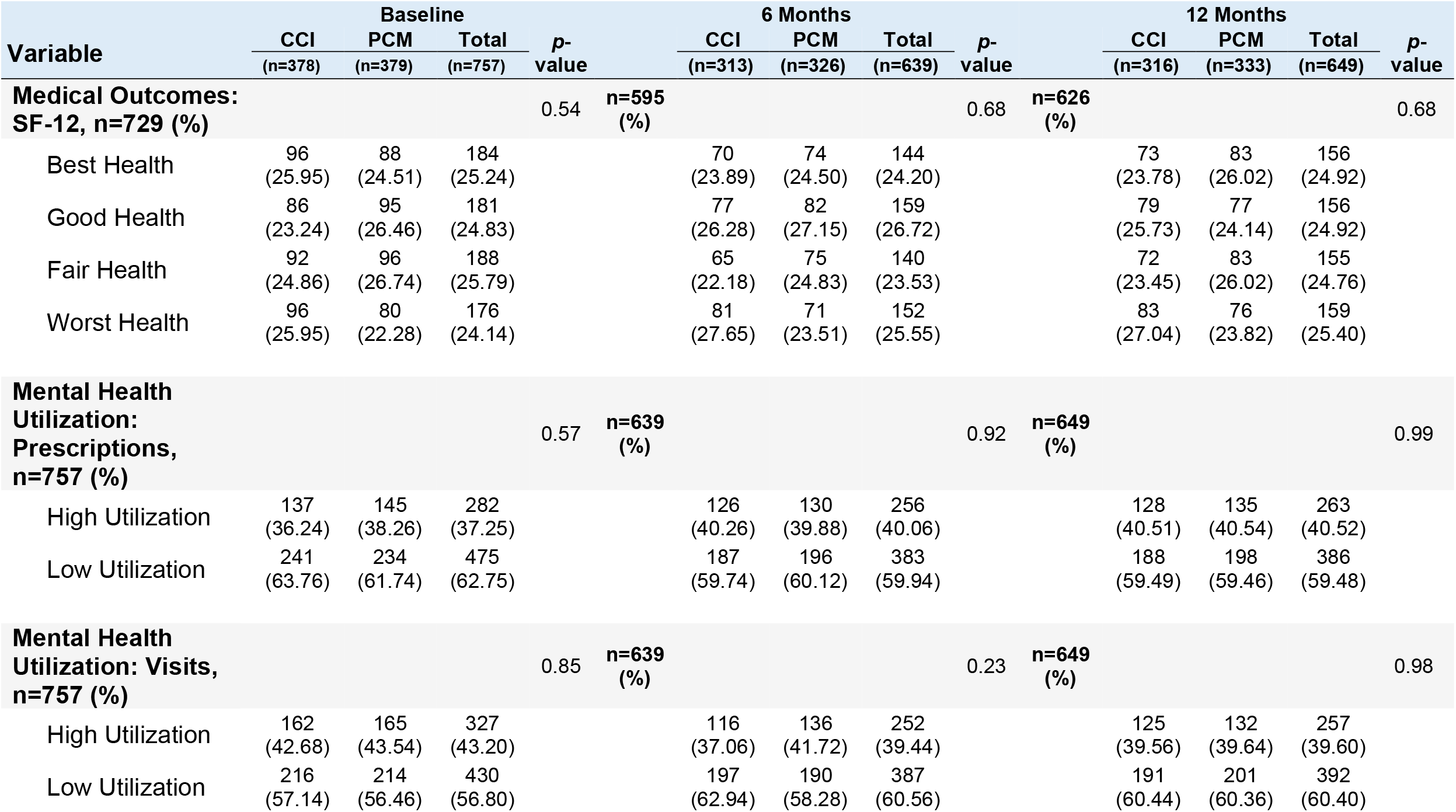
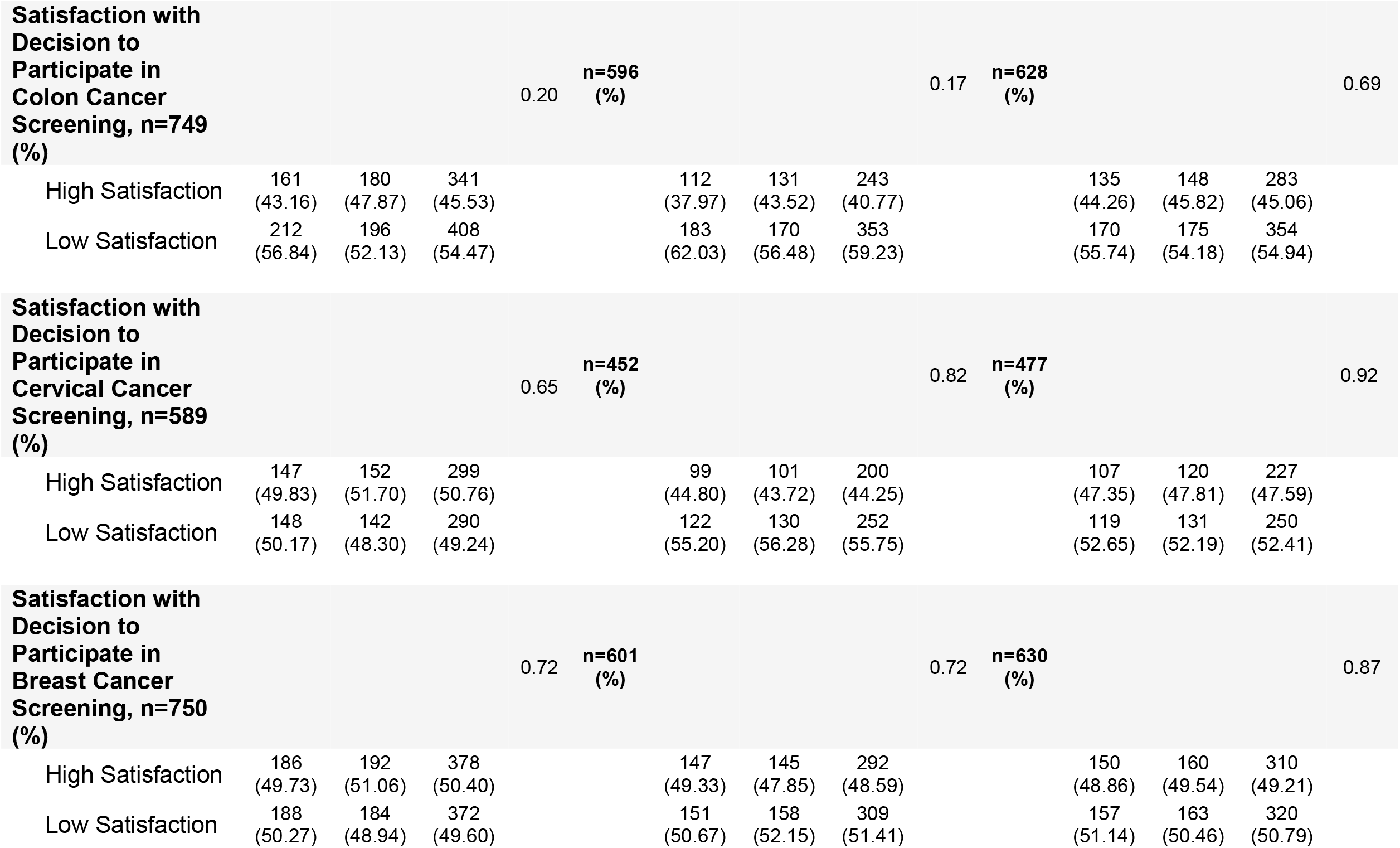
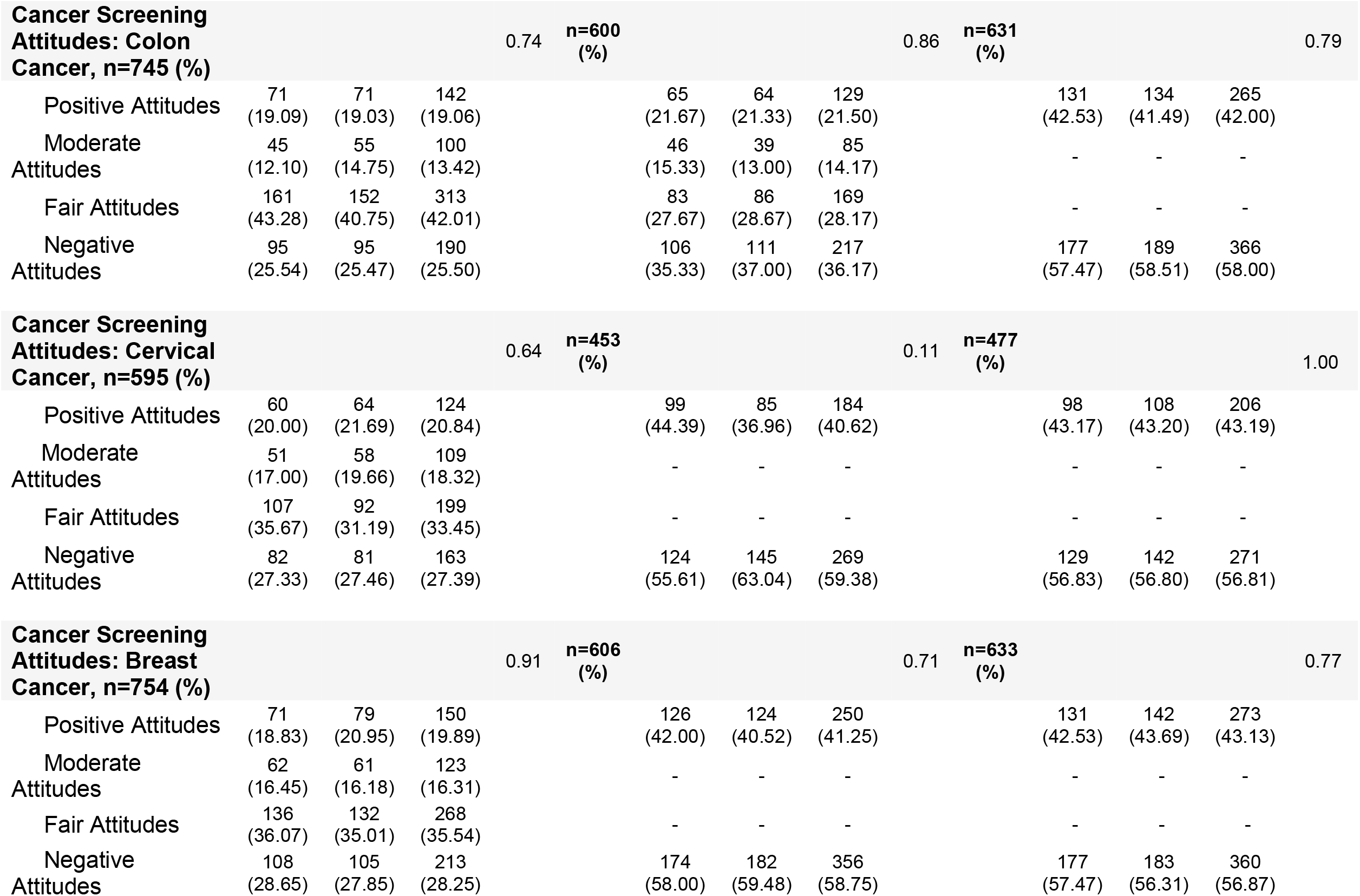
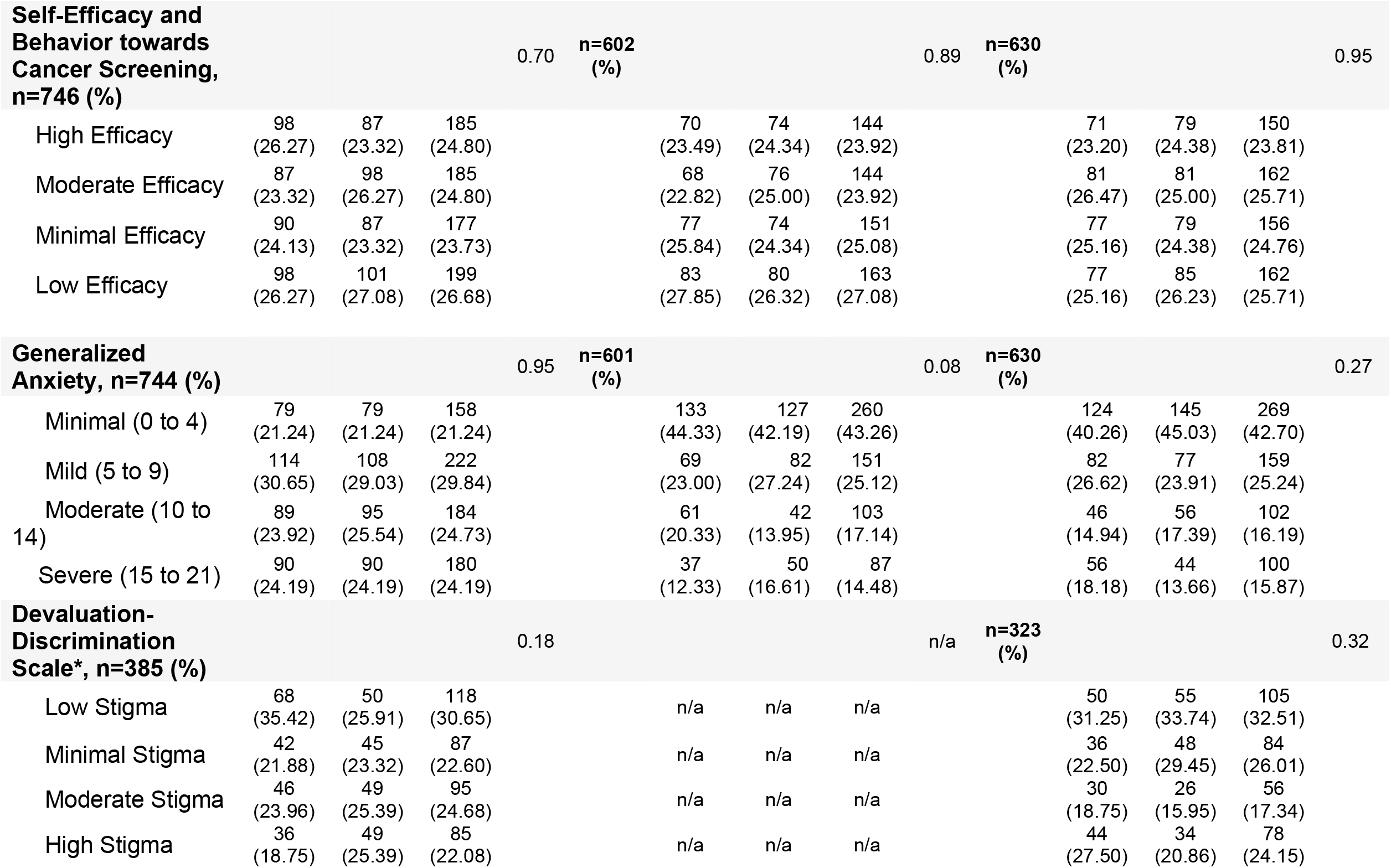
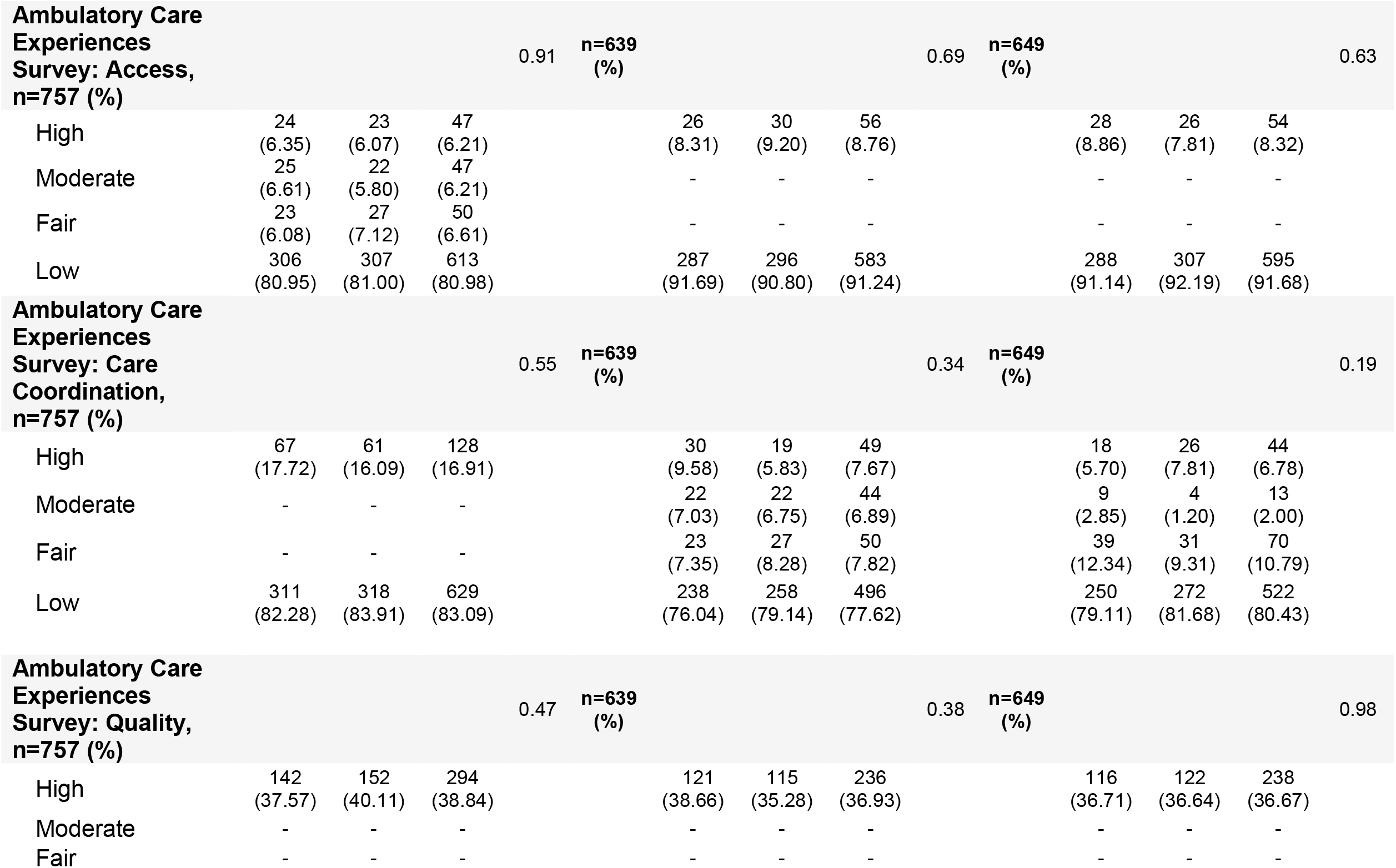
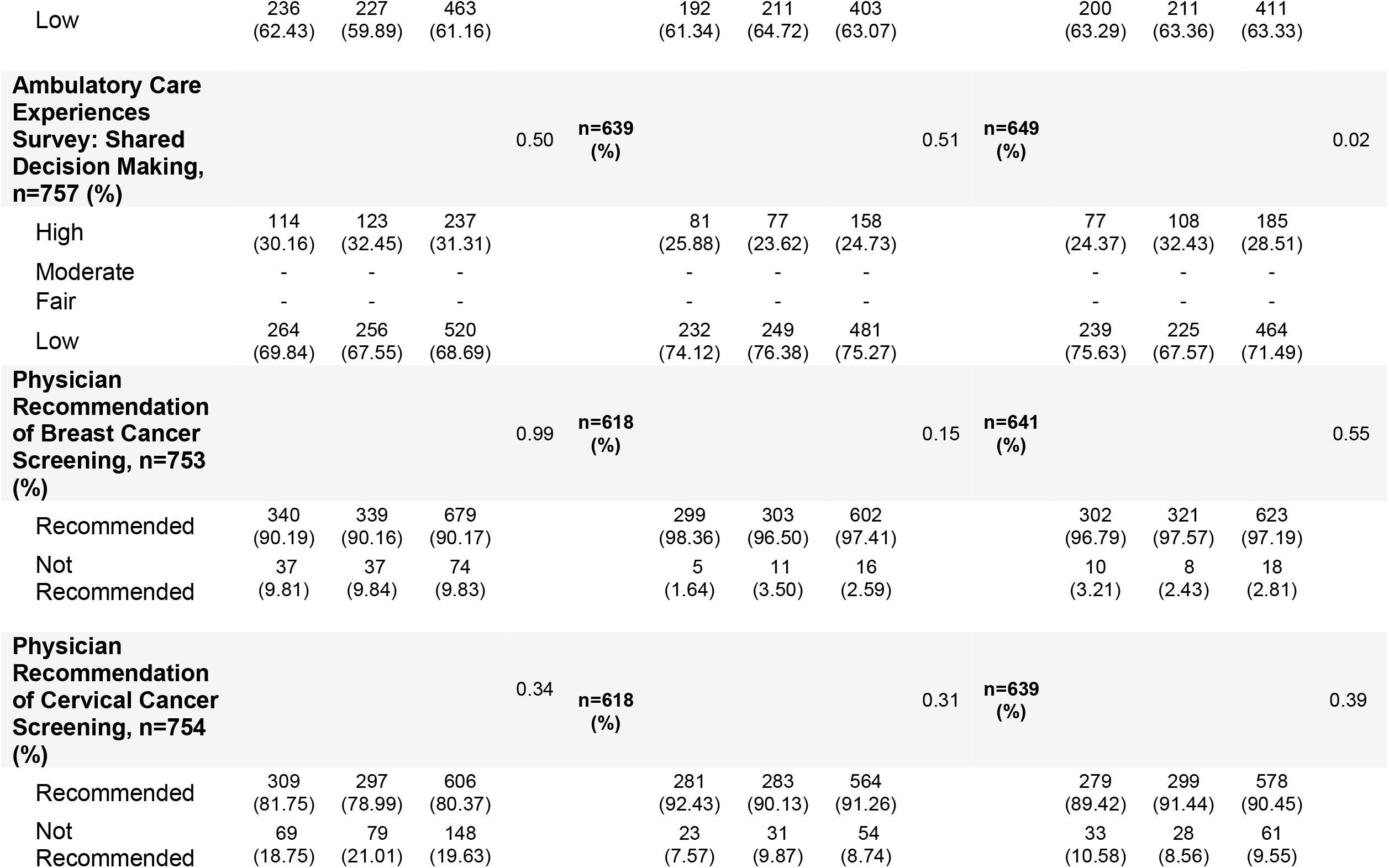
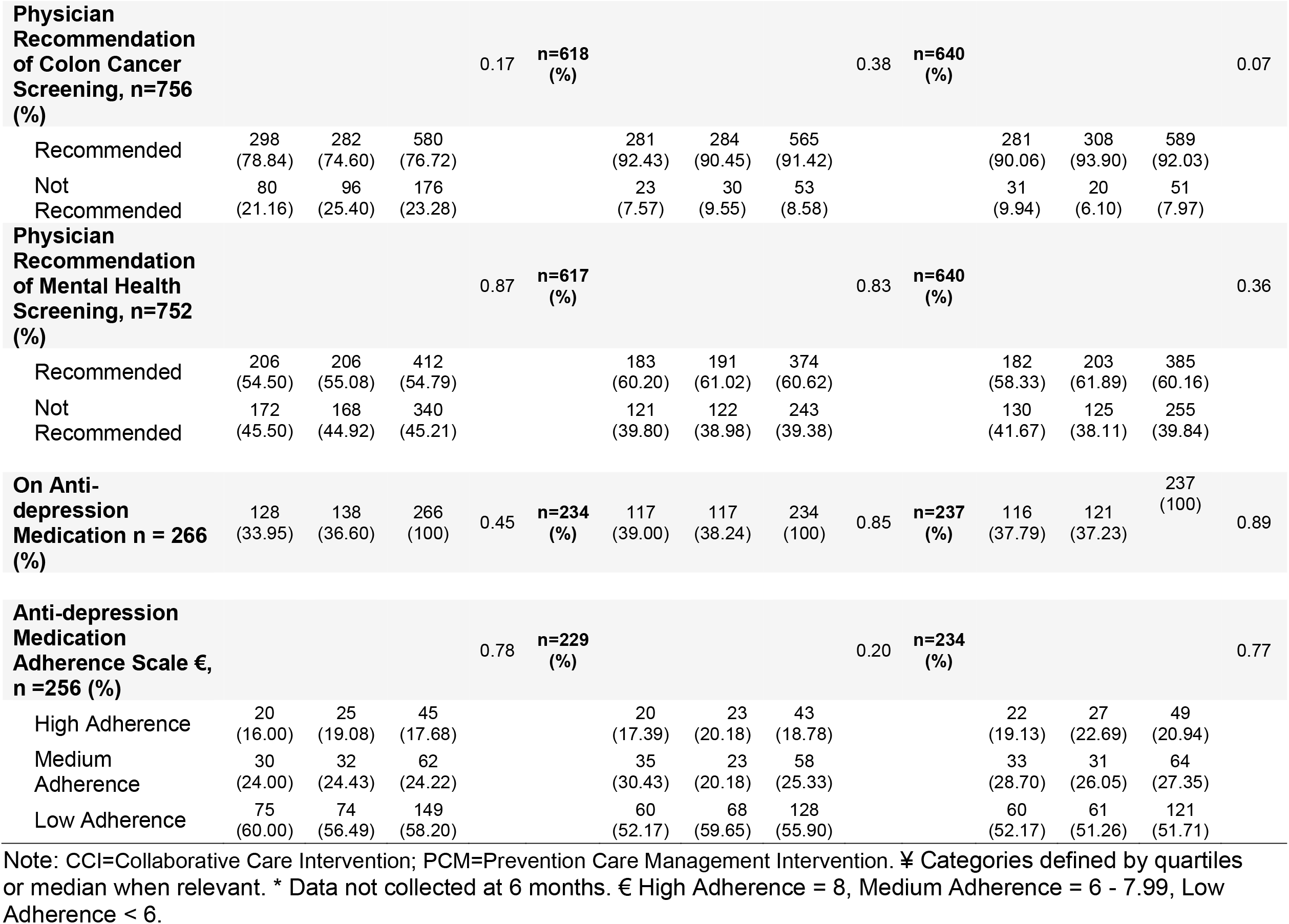
Patient-reported outcomes by treatment condition at baseline, 6-month, and 12-month follow-up **¥ £**

**APPENDIX Table A6.**
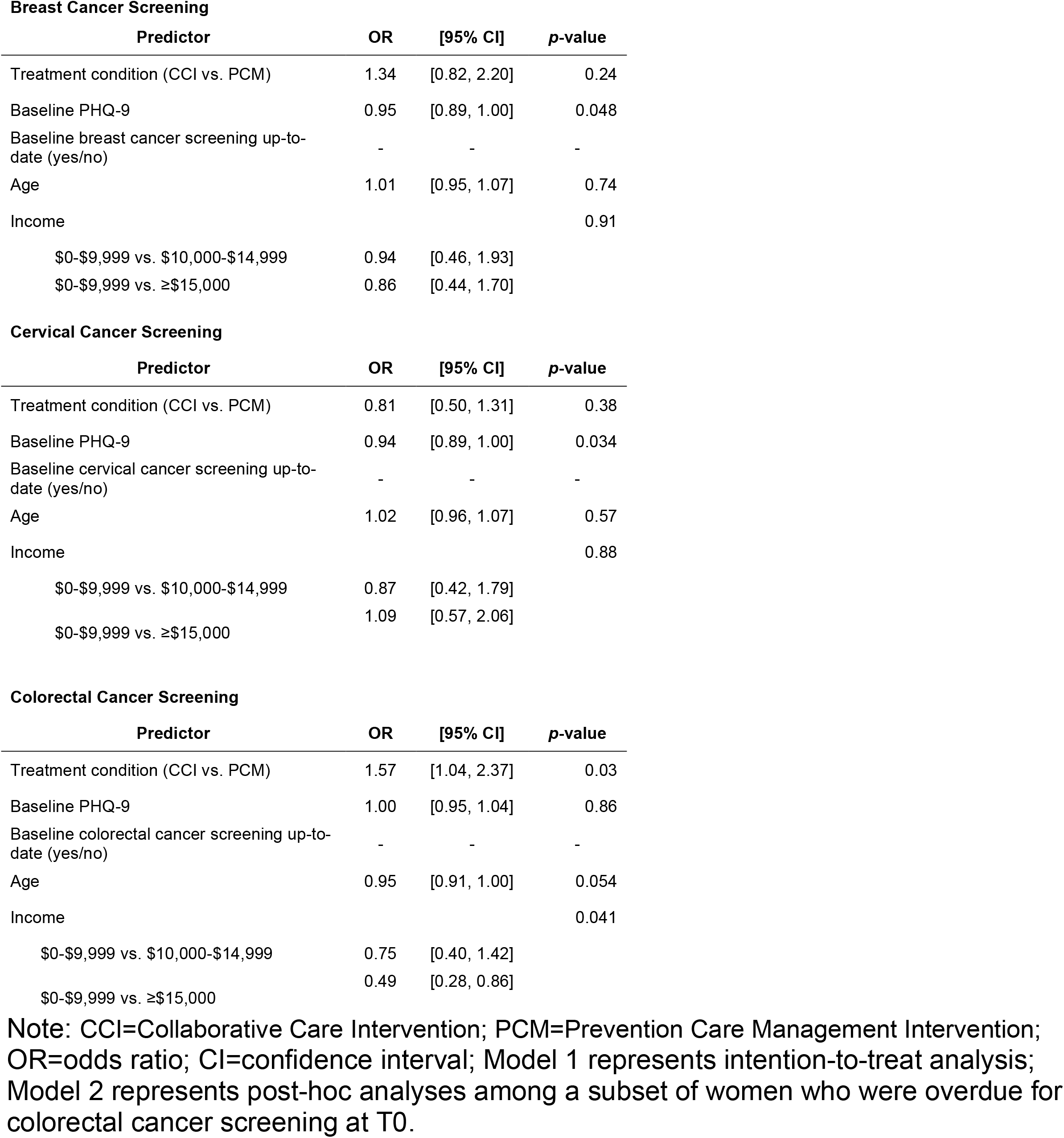
Logistic regression models of cancer screenings post-intervention (12 months)

**APPENDIX Table A7.** Logistic regression model of screening status for all cancers post-intervention (12 months)

| Predictor | OR | [95% CI] | p-value |
| --- | --- | --- | --- |
| Treatment condition (CCI vs. PCM) | 1.09 | [0.84, 1.43] | 0.51 |
| Baseline PHQ-9 | 0.97 | [0.94, 1.00] | 0.061 |
| Baseline number of cancer screenings up-to-date |  |  | <0.001 |
| 0 vs. 1 | 2.22 | [1.58, 3.11] |  |
| 0 vs. 2 | 5.24 | [3.72, 7.36] |  |
| Age | 0.98 | [0.95, 1.02] | 0.30 |
| Income |  |  | 0.40 |
| \$0-\$9,999 vs. \$10,000-\$14,999 | 1.04 | [0.71, 1.51] | |
| \$0-\$9,999 vs. ≥\$15,000 | 0.80 | [0.56, 1.13] | |

Note: CCI=Collaborative Care Intervention; PCM=Prevention Care Management Intervention; OR=odds ratio; CI=confidence interval.

**APPENDIX Table A8.**
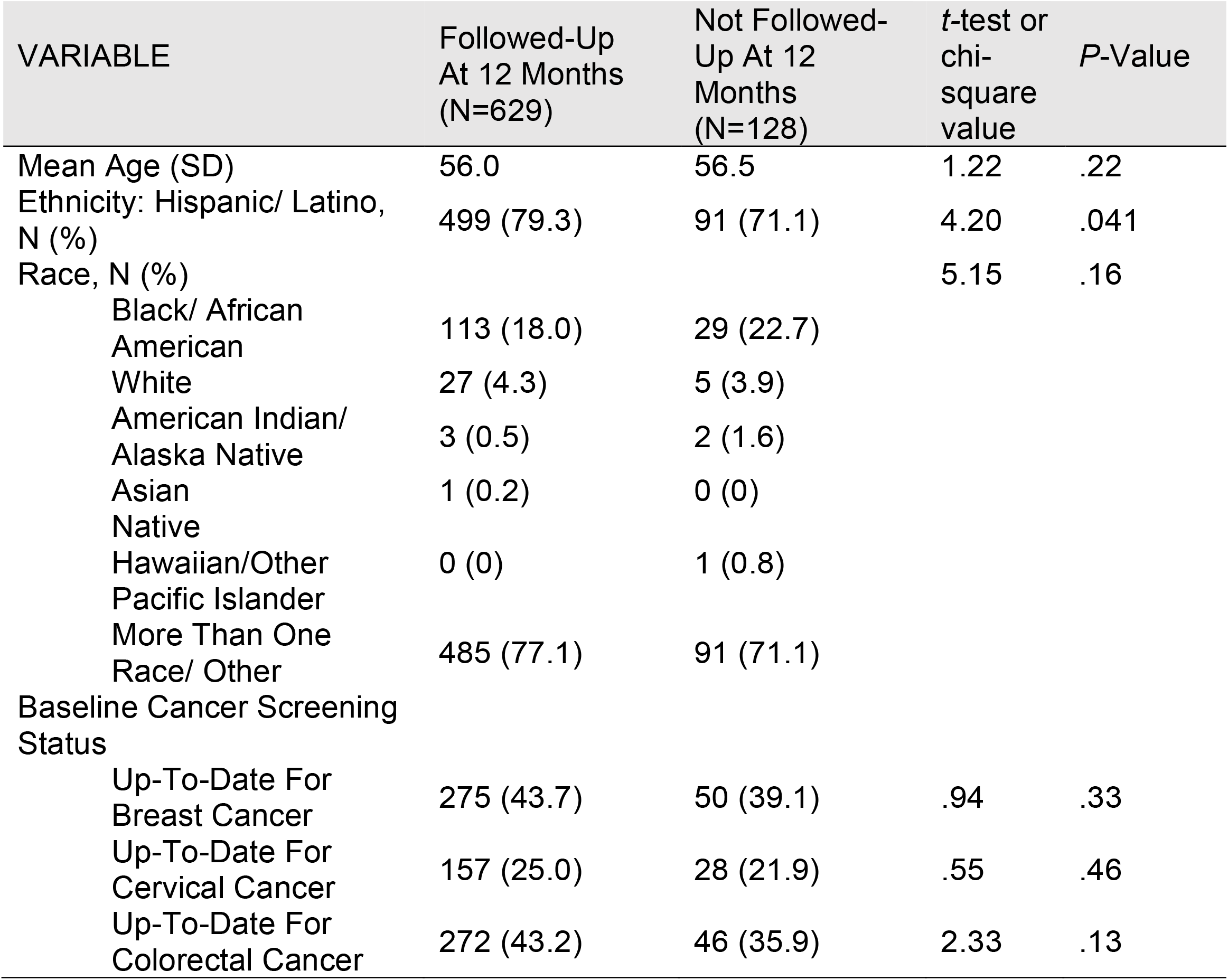
Characteristics of participants who were followed-up vs. not followed-up for depression measures

**APPENDIX Table A9.** Logistic regression model of depression reduction and depression remission post-intervention (12 months)

| Predictor | Depression Improvement |  |  | Depression Remission |  |  |
| --- | --- | --- | --- | --- | --- | --- |
|  | OR | [95% CI] | p-value | OR | [95% CI] | p-value |
| Treatment condition (CCI vs. PCM) | 1.33 | [0.91, 1.93] | 0.14 | 0.88 | [0.63, 1.25] | 0.48 |
| Baseline PHQ-9 | 1.08 | [1.03, 1.13] | 0.002 | 0.88 | [0.84, 0.92] | <0.001 |
| Baseline number of cancer screenings up-to-date |  |  | 0.63 |  |  | 0.23 |
| 0 vs. 1 | 0.80 | [0.49, 1.29] |  | 0.69 | [0.45, 1.07] |  |
| 0 vs. 2 | 0.92 | [0.57, 1.48] |  | 0.90 | [0.59, 1.36] |  |
| Age | 1.01 | [0.97, 1.06] | 0.66 | 1.03 | [0.99, 1.08] | 0.13 |
| Income |  |  | 0.061 |  |  | 0.037 |
| \$0-\$9,999 vs. \$10,000-\$14,999 | 1.55 | [0.88, 2.73] | | 1.72 | [1.07, 2.77] | |
| \$0-\$9,999 vs. ≥\$15,000 | 1.78 | [1.03, 3.07] | | 1.48 | [0.94, 2.32] | |

Note: Note: CCI=Collaborative Care Intervention; PCM=Prevention Care Management Intervention; OR=odds ratio; CI=confidence interval.

